# Whole blood DNA methylation changes are associated with anti-TNF drug concentration in patients with Crohn’s disease

**DOI:** 10.1101/2023.03.22.23287574

**Authors:** Simeng Lin, Eilis Hannon, Mark Reppell, Jeffrey F. Waring, Nizar Smaoui, Valerie Pivorunas, Heath Guay, Neil Chanchlani, Claire Bewshea, Benjamin Y H Bai, Nicholas A Kennedy, James R Goodhand, Jonathan Mill, Tariq Ahmad, PANTS Consortium

## Abstract

**Background and Aims:** Anti-TNF treatment failure in patients with inflammatory bowel disease (IBD) is common and frequently related to low drug concentrations.

In order to identify patients who may benefit from dose optimisation at the outset of anti-TNF therapy, we sought to define epigenetic biomarkers in whole blood at baseline associated with anti-TNF drug concentrations at week 14.

**Methods:** DNA methylation from 1,104 whole blood samples from the Personalised Anti-TNF Therapy in Crohn’s disease (PANTS) study were assessed using the Illumina EPIC Beadchip at baseline, weeks 14, 30 and 54. We compared DNA methylation profiles in anti-TNF-treated patients who experienced primary non-response at week 14 and were not in remission at week 30 or 54 (infliximab n = 99, adalimumab n = 94) with patients who responded at week 14 and were in remission at week 30 or 54 (infliximab n = 99, adalimumab n = 93).

**Results:** Overall, between baseline and week 14, we observed 4,999 differentially methylated probes (DMPs) annotated to 2376 genes following anti-TNF treatment. Pathway analysis identified 108 significant gene ontology terms enriched in biological processes related to immune system processes and responses.

Epigenome-wide association (EWAS) analysis identified 323 DMPs annotated to 210 genes at baseline associated with higher anti-TNF drug concentrations at week 14. Of these, 125 DMPs demonstrated shared associations with other common traits (proportion of shared CpGs compared to DMPs) including body mass index (23.2%), followed by CRP (11.5%), smoking (7.4%), alcohol consumption per day (7.1%) and IBD type (6.8%). EWAS of primary non-response to anti-TNF identified 20 DMPs that were associated with both anti-TNF drug concentration and primary non-response to anti-TNF with a strong correlation of the coefficients (Spearman’s rho = −0.94, p < 0.001).

**Conclusion:** Baseline DNA methylation profiles may be used as a predictor for anti-TNF drug concentration at week 14 to identify patients who may benefit from dose optimisation at the outset of anti-TNF therapy.

## Introduction

Anti-TNF therapies remain the most effective treatment to induce and maintain remission in patients with Crohn’s disease^1, 2^. Successful treatment leads to mucosal healing, reduced surgeries, and improvements in quality of life^3^. Unfortunately, anti-TNF treatment failure is common, with a quarter of patients experiencing primary non-response, and one-third of initial responders losing response by the end of the first year^4^.

In the Personalised Anti-TNF Therapy in Crohn’s Disease study (PANTS), whilst loss of response was associated with the formation of anti-drug antibodies that were predicted by carriage of the HLA-DQA1*05 haplotype and mitigated by concomitant immunomodulator use, the only modifiable factor associated with primary non-response at week 14 was low anti-TNF drug concentration^5, 6^.

In this regard early dose optimisation reportedly improves anti-TNF response rates^7, 8^. Whilst the biology of non-response is complex, an ability to predict primary non-response may inform treatment choice and identify individuals who may benefit from dose optimisation during induction therapy.

Heterogeneity of response to anti-TNF therapies has led to a drive to understand the molecular mechanisms underlying treatment failure in anti-TNF therapy. Increased mucosal expression of oncostatin M (*OSM*)^9, 10^ and triggering receptor expressed on myeloid cells (*TREM-1*)^11, 12^ have been identified as potential biomarkers predicting non-response to anti-TNF treatment. Drawing conclusions across studies is difficult, however, due to differences in study design, improvements in experimental and computational methods through time and critically, confounding by cellular heterogeneity with contradictory results between whole blood and intestinal biopsies^13^. Clinical translation of tissue biomarkers has also further been limited by accessibility and processing costs^14^. DNA methylation, an epigenetic modification to DNA, can influence gene expression via disruption of transcription factor binding and recruitment of methyl-binding proteins that initiate chromatin compaction and gene silencing^15, 16^. Despite being traditionally regarded as a mechanism of transcriptional repression, DNA methylation is actually associated with both increased and decreased gene expression^17^, and other genomic functions including alternative splicing and promoter usage^18^. DNA methylation can be influenced by both genetic^19^ and environmental factors^20^, changing with age^21^ and exposures such as cigarette smoking^22^. The development of standardised assays for quantifying DNA methylation across the genome at single-base resolution in large numbers of samples has enabled researchers to perform epigenome-wide association studies (EWAS) aimed at identifying methylomic variation associated with exposures and traits^23^. EWAS analyses are inherently more complex to design and interpret than genetic association studies; the dynamic nature of epigenetic processes means that a range of potentially important confounding factors (including tissue or cell type, age, sex, and lifestyle exposures) need to be considered in between-group comparisons^24^.

Previous studies of DNA methylation using whole blood or individual purified cell types have identified differentially methylated positions (DMPs) between patients with active and inactive IBD and healthy controls^25, 26^. Pharmacoepigenomics is the application of epigenetics to understand interindividual differences in the response to therapeutic drugs^27^. DNA methylation sites from whole blood have been identified as effective biomarkers predicting treatment response to methotrexate and anti-TNF in patients with rheumatoid arthritis^28, 29^.

In this study we used a powerful intra-individual study design to identify changes in DNA methylation associated with anti-TNF drug treatment, profiling 385 patients at baseline and weeks 14, 30 and 54 post treatment initiation. In order to identify patients who may benefit from dose optimisation at the outset of anti-TNF therapy, we also sought to define epigenetic biomarkers in whole blood at baseline associated with anti-TNF drug concentrations at week 14. We identify widespread differences in DNA methylation induced by anti-TNF drug treatment and show that baseline DNA methylation profiles can predict anti-TNF drug concentration at week 14.

## Methods

### Study design

The PANTS study is a UK wide, multicentre, prospective observational cohort reporting the treatment failure rates of the anti-TNF drugs infliximab (originator, Remicade [Merck Sharp & Dohme, Hertfordshire, UK] and biosimilar, CTP13 [Celltrion, Incheon, South Korea]), and adalimumab (Humira [AbbVie, Cambridge, MA]) in 1610 anti–TNF-naïve patients with Crohn’s disease. (Supplementary Table 1)

Patients were recruited between February 2013 and June 2016 at the time of first anti-TNF exposure and studied for 12 months or until drug withdrawal. Eligible patients were aged ≥ 6 years with objective evidence of active luminal Crohn’s disease involving the colon and/or small intestine. Exclusion criteria included prior exposure to, or contraindications for the use of, anti-TNF therapy. The choice of anti-TNF was at the discretion of the treating physician and prescribed according to the licensed dosing schedule. Study visits were scheduled at first dose, week 14, and at weeks 30 and 54. Additional visits were planned for infliximab-treated patients at each infusion and for both groups at treatment failure or exit.

Treatment failure endpoints were primary non-response at week 14, non-remission at week 54, and adverse events leading to drug withdrawal.

We used composite endpoints using the Harvey Bradshaw Index (HBI) in adults and the short paediatric Crohn’s disease activity index (sPCDAI) in children, corticosteroid use, and CRP to define primary non-response (Supplementary Figure 1). Remission was defined as CRP of ≤3 mg/L and HBI of ≤4 points (sPCDAI ≤15 in children), without corticosteroid therapy or exit for treatment failure.

Variables recorded at baseline were patient demographics (age, sex, ethnicity, comorbidities, height and weight and smoking status) and IBD phenotype and its treatments (age at diagnosis, disease duration, Montreal classification, prior medical and drug history, and previous Crohn’s disease-related surgeries). At every visit, disease activity score, weight, current therapy and adverse events were recorded.

Blood and stool samples were collected at each visit and processed through the central laboratory at the Royal Devon & Exeter NHS Foundation Trust (https://www.exeterlaboratory.com/) for haemoglobin, white cell count, platelets, serum albumin, CRP, anti-TNF drug and anti-drug antibody concentrations, and faecal calprotectin, respectively.

### DNA methylation processing

Genomic DNA was extracted from peripheral whole blood using the Qiagen Qiasymphony DNA DSP midi kit (Qiagen, Ca, USA). Following sodium bisulfite conversion with the Zymo Research EZ-DNA Methylation kit (Zymo Research, CA USA), DNA methylation was quantified across the genome using the Illumina Infinium HumanMethylationEPIC (EPIC) BeadChip (Illumina Inc, CA, USA). To negate any methodological batch effects, individuals were randomised across experimental batches and samples from the same individual were processed together across all experimental stages.

### Data pre-processing and quality control checks

Raw Illumina EPIC data were imported into R (version 3.6.0) using the *bigmelon* package (v1.12.0)^30^. Quality control (QC) checks were performed using the *bigmelon* (v1.12.0)^30^ and *minfi* (v1.32.0)^31^ R packages. They included the following steps: we first removed samples by 1) checking median methylated and unmethylated signal intensities and excluding samples with low intensities (<500) (3 samples excluded), 2) assessing bisulphite conversion efficacy of each sample and excluding samples with a conversion rate < 80% (9 samples excluded), 3) using the 59 single nucleotide polymorphism (SNP) probes present on the EPIC array to confirm all matched samples from the same individual were genetically identical and to check for sample switches or duplications (12 samples excluded), 4) comparing intensity values from probes located on the X and Y chromosomes to autosomes to identify sex mismatches (10 samples excluded), 5) visually inspecting the first six principal components and excluding outliers (none identified), 6) using the pfilter function in the *bigmelon* (v1.12.0) package to exclude samples where >1% of probes had a detection p-value >0.05 (none identified). We subsequently removed probes by 7) using the pfilter function in the *bigmelon* (v1.12.0) package to exclude probes with a bead count <3 or 1% of samples with a detection p-value > 0.05 (8313 probes) and 8) removal of cross-hybridising probes and those containing a SNP (73,239 probes). Following QC, quantile normalisation was carried out and 784,105 probes were taken forwards for analysis after exclusion of probes on the Y chromosome.

### Sample size and statistical methods

Sample size calculations from the whole PANTS cohort have been reported previously^5^. For this analysis, we selected whole blood samples from a subset of 385 participants treated with infliximab and adalimumab aged >16 years, with a baseline CRP ≥4 mg/L and/or calprotectin >100 µg/g who experienced primary non-response at week 14 and non-remission at week 30 or 54 (n = 99 and 94, respectively), and an equal number of participants as a comparator group who were classified with primary response at week 14 and subsequent remission (n = 99 and 93, respectively) for DNA methylation profiling.

Statistical analyses were undertaken in R 4.1.3 (R Foundation for Statistical Computing, Vienna, Austria). We included patients with missing clinical variables in analyses for which they had the necessary data and have specified the sample size for each variable. Continuous data are reported as median and interquartile range, and discrete data as numbers and percentages. Fisher’s exact and Mann-Whitney U tests were used to identify differences in baseline characteristics between infliximab-treated and adalimumab-treated patients. Comparative tests were two-tailed and p value < 0.05 were considered significant unless otherwise stated.

DNA methylation was analysed using beta values, the ratio of methylated intensity to the overall intensity at each CpG site, which represents the proportion methylation at each site. Because they influence methylation, *a priori,* we sought to define DNA methylation changes through the course of the study due to ageing, cigarette smoking and cell composition. Smoking scores were calculated using a weighted sum approach based on previously published smoking-associated methylation probes^32^. Epigenetic age was predicted using 353 CpG sites as described by Horvath^21^. Individual cell proportions of CD4+T cells, CD8+T cells, B cells, granulocytes, and monocytes in each whole blood sample were estimated using the Houseman reference-based algorithm implemented with functions in *minfi* (v1.32.0)^31^ package. Linear mixed effects models, including time on anti-TNF (study visits in weeks) as a fixed effect and modelling individual participants with a random intercept, were used to determine if epigenetic age, smoking behaviour or cell composition were associated with anti-TNF treatment.

EWAS analyses of anti-TNF treatment, anti-TNF drug concentration and primary non-response were conducted using linear mixed effects models in this within-subject study, where anti-TNF type and cell proportions were adjusted for as fixed effects while a random effect (random intercept) was used to capture the individual level effects. Study visits in weeks, reflecting the duration of anti-TNF treatment, was included as an interaction term in the model. Patients treated with infliximab and adalimumab were analysed together to increase the power to detect shared effects. An empirically-derived p value < 9 x 10^-8^ was considered significant to control for multiple testing^33^. Pathway analysis with annotations to Gene Ontology (GO) terms was performed using the *gometh ()* function in *missMethyl* (v1.28.0)^34^ package, which controls for bias arising due to multiple genes being annotated to a single CpG, and multiple CpGs annotated to a single gene. DMPs were searched in the EWAS catalog^35^ (http://www.ewascatalog.org/, assessed on 15/12/2022) to look for associations with other common traits. A false discovery rate (FDR) of < 0.05 was considered significant for pathway analysis and associations in the EWAS catalog. We sought overlapping DMPs associated with drug levels and primary non-response and correlation of coefficients was determined using Spearman’s test.

### Ethical and role of the funding source

The sponsor of the study was the Royal Devon and Exeter NHS Foundation Trust. The South West Research Ethics committee approved the study (REC Reference: 12/SW/0323) in January 2013. The funders of this study had no role in study design, data collection, data analysis, data interpretation, or writing of the report. The corresponding author had full access to all the data in the study and had final responsibility for the decision to submit for publication.

## Results

### Summary of PANTS DNA methylation dataset

DNA methylation was quantified across the genome in 1,104 whole blood DNA samples from 385 individuals across four study visits (baseline, week 14, week 30, week 54) from the PANTS cohort. Following a standard quality control pipeline (see Methods), our final dataset included 784,105 DNA methylation sites quantified in 1,062 samples from 385 participants (Supplementary Figure 1). 87 participants provided samples at all four study visits and the median number of samples per participant was 3 (interquartile range [IQR] 2 – 3) (Supplementary Table 2).

Overall, 51.7% (199/385) of participants were female, with a median age of 35.7 years (IQR 26.3 - 50.3). 21.2% (81/382) of participants were current smokers and 30.6% (117/382) were former smokers. The median disease duration was 2.2 years [IQR 0.6 - 9.6] and 50.9% (196/385) and 35.3% (136/385) of participants were treated with a concomitant immunomodulator and steroids at baseline, respectively. In total, 51.4% (198/385) of participants were treated with infliximab and 48.6% (187/385) with adalimumab (Table 1). Median infliximab (3.30 mg/L vs 8.09 mg/L, p < 0.001) and adalimumab (7.70 mg/L vs. 13.35 mg/L, p < 0.001) drug concentrations at week 14 were lower in patients who experienced primary non-response, as previously observed in the wider cohort^5^. (Supplementary Figure 2).

**Table 1:**
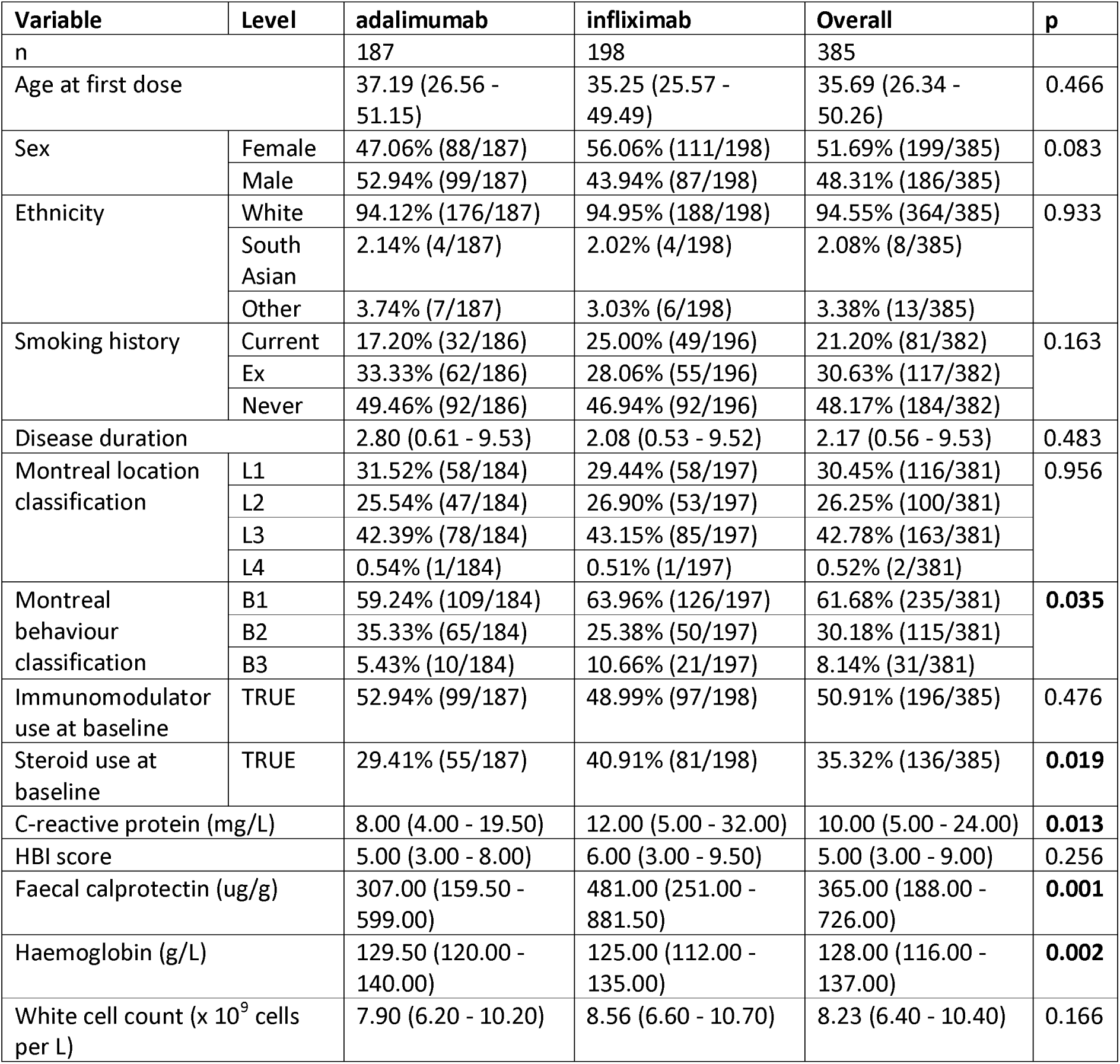
Characteristics at baseline of participants stratified by type of anti-TNF

| Variable | Level | adalimumab | infliximab | Overall | p |
| --- | --- | --- | --- | --- | --- |
| n |  | 187 | 198 | 385 |  |
| Age at first dose |  | 37.19 (26.56 - 51.15) | 35.25 (25.57 - 49.49) | 35.69 (26.34 - 50.26) | 0.466 |
| Sex | Female | 47.06% (88/187) | 56.06% (111/198) | 51.69% (199/385) | 0.083 |
|  | Male | 52.94% (99/187) | 43.94% (87/198) | 48.31% (186/385) |  |
| Ethnicity | White | 94.12% (176/187) | 94.95% (188/198) | 94.55% (364/385) | 0.933 |
|  | South Asian | 2.14% (4/187) | 2.02% (4/198) | 2.08% (8/385) |  |
|  | Other | 3.74% (7/187) | 3.03% (6/198) | 3.38% (13/385) |  |
| Smoking history | Current | 17.20% (32/186) | 25.00% (49/196) | 21.20% (81/382) | 0.163 |
|  | Ex | 33.33% (62/186) | 28.06% (55/196) | 30.63% (117/382) |  |
|  | Never | 49.46% (92/186) | 46.94% (92/196) | 48.17% (184/382) |  |
| Disease duration |  | 2.80 (0.61 - 9.53) | 2.08 (0.53 - 9.52) | 2.17 (0.56 - 9.53) | 0.483 |
| Montreal location classification | L1 | 31.52% (58/184) | 29.44% (58/197) | 30.45% (116/381) | 0.956 |
|  | L2 | 25.54% (47/184) | 26.90% (53/197) | 26.25% (100/381) |  |
|  | L3 | 42.39% (78/184) | 43.15% (85/197) | 42.78% (163/381) |  |
|  | L4 | 0.54% (1/184) | 0.51% (1/197) | 0.52% (2/381) |  |
| Montreal behaviour classification | B1 | 59.24% (109/184) | 63.96% (126/197) | 61.68% (235/381) | 0.035 |
|  | B2 | 35.33% (65/184) | 25.38% (50/197) | 30.18% (115/381) |  |
|  | B3 | 5.43% (10/184) | 10.66% (21/197) | 8.14% (31/381) |  |
| Immunomodulator use at baseline | TRUE | 52.94% (99/187) | 48.99% (97/198) | 50.91% (196/385) | 0.476 |
| Steroid use at baseline | TRUE | 29.41% (55/187) | 40.91% (81/198) | 35.32% (136/385) | 0.019 |
| C-reactive protein (mg/L) |  | 8.00 (4.00 - 19.50) | 12.00 (5.00 - 32.00) | 10.00 (5.00 - 24.00) | 0.013 |
| HBI score |  | 5.00 (3.00 - 8.00) | 6.00 (3.00 - 9.50) | 5.00 (3.00 - 9.00) | 0.256 |
| Faecal calprotectin (ug/g) |  | 307.00 (159.50 - 599.00) | 481.00 (251.00 - 881.50) | 365.00 (188.00 - 726.00) | 0.001 |
| Haemoglobin (g/L) |  | 129.50 (120.00 - 140.00) | 125.00 (112.00 - 135.00) | 128.00 (116.00 - 137.00) | 0.002 |
| White cell count (x 10<sup>9</sup> cells per L) |  | 7.90 (6.20 - 10.20) | 8.56 (6.60 - 10.70) | 8.23 (6.40 - 10.40) | 0.166 |

### Anti-TNF treatment is associated with altered blood cell proportions using measures derived from DNA methylation data

A number of robust statistical classifiers have been developed to derive estimates of environmental exposures such as tobacco smoking^32^, biological age^21^ and the proportion of different blood cell types^36^ from whole blood DNA methylation data.

As expected, current and former tobacco smokers had a higher DNA methylation-derived smoking score at baseline (former smokers 0.2 [IQR −2.0 - 4.1], p < 0.001 and current smokers 6.6 [IQR 3.3 - 9.5], p < 0.001) when compared to never smokers (−2.3 [IQR −3.8 - −1.0]). Over the duration of the study, DNA methylation smoking score increased (effect size per week 0.019, p < 0.001). When compared to current smokers, the trajectory of DNA methylation smoking score changed significantly in former (effect size per week −0.010, p = 0.003), but not current smokers (effect size per week −0.004, p = 0.262) (Supplementary Figure 3) When stratified by response to anti-TNF treatment, following anti-TNF treatment, there was no difference in the trajectory of smoking scores of current smokers (effect size per week - 0.006, p = 0.433) or former smokers (effect size per week - 0.0001, p = 0.986) between those who experienced primary non-response compared to those who did not.

The epigenetic age of participants measured using the Horvath multi-tissue clock was highly correlated with chronological age of participants at study entry (r = 0.95, p < 0.001). Over the course of the study following anti-TNF treatment, epigenetic age changed with time (effect size per week 0.002, p < 0.001). When stratified by response to anti-TNF treatment, however, there was no difference in the trajectory of change in epigenetic age (effect size per week 0.003, p = 0.71). (Supplementary Figure 4)

To understand the immune cell changes following anti-TNF treatment, cell proportion estimates were derived from DNA methylation data. Over time, following anti-TNF treatment, there was a significant increase in the derived proportions of CD4 T cells (effect size per week 0.0013, p < 0.001), CD8 T cells (effect size per week 0.0005, p < 0.001), B cells (effect size per week 0.0004, p < 0.001) and NK cells (effect size per week 0.0001, p = 0.015). (Figure 1) In contrast, the proportion of monocytes (effect size per week −0.0001, p = 0.025) and granulocytes (effect size per week −0.0023, p < 0.001) decreased significantly. In patients who experienced primary non-response, the increase in proportion of B cells (effect size per week −0.0002, p < 0.001) and CD4 T cells (effect size per week −0.0004, p = 0.048) were less marked over time when compared to those who responded. There was no difference in the change of proportion of granulocytes (effect size per week 0.0002, p = 0.571).

**Figure 1:**
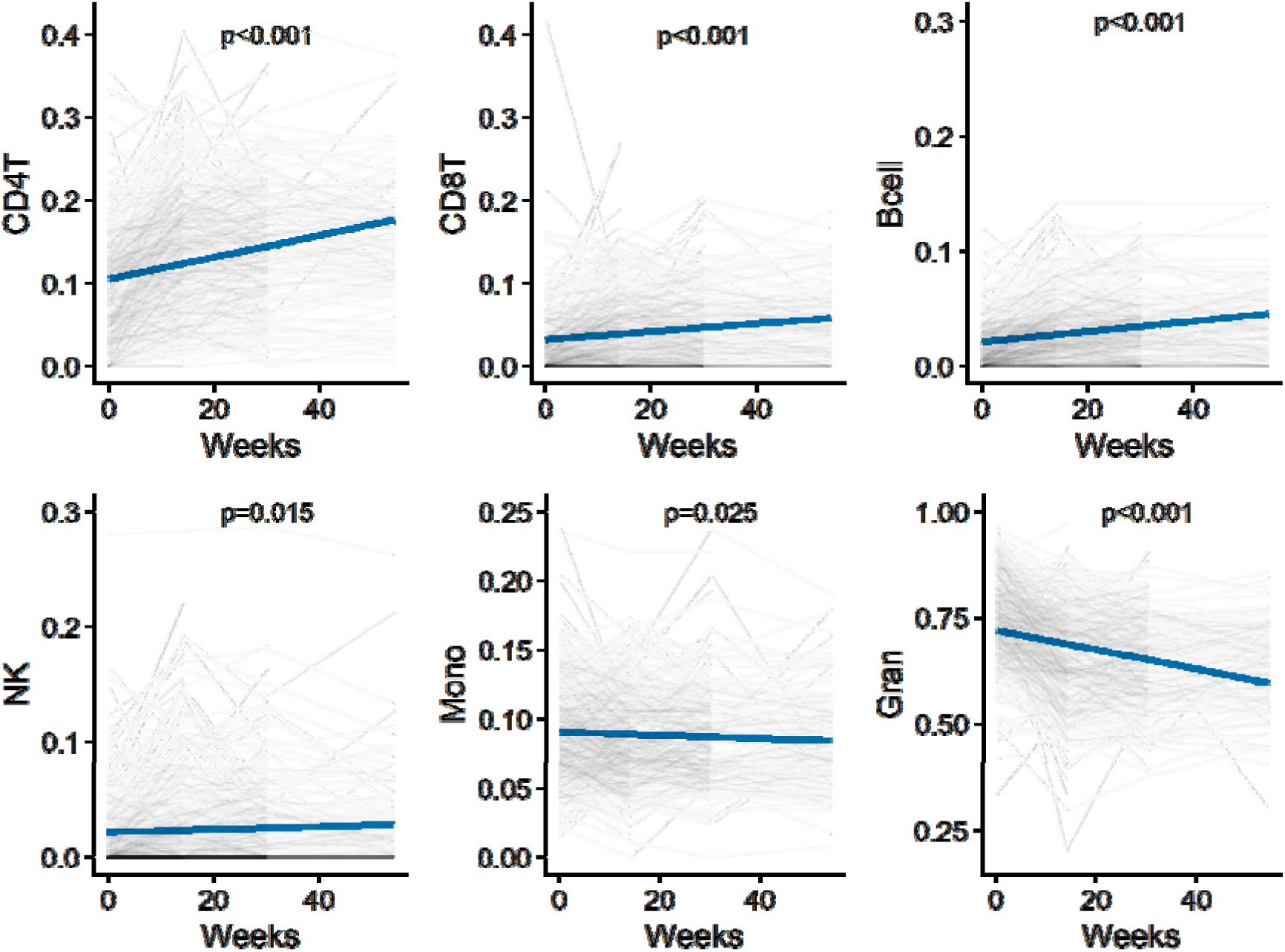
Change in derived cell proportions following treatment with anti-TNF. Predicted derived cell proportions over time estimated from the regression analysis is represented in solid blue lines, and observed cell proportions in faded lines. P value represents the change in individual cell proportions over time.

### Changes in biological processes of the immune pathways occur following anti-TNF treatment

Across all patients, 4,999 DMPs (p < 9 x 10^-8^) annotated to 2,376 unique genes were associated with anti-TNF treatment (infliximab or adalimumab) regardless of response (Table 2, Figure 2). These DMPs were significantly enriched for sites becoming hypomethylated over time (63.5% [3,176/4,999], p < 0.001). Of treatment-associated DMPs annotated to genes (n = 3,504 [70.1%]), the majority were located in the gene body (67.1% [2,351/3,504]) (Supplementary Figure 5) representing a significant enrichment compared to the background distribution of probes on the EPIC array (67.1% vs 29.5%, p < 0.001). The top-ranked DMP associated with anti-TNF treatment was cg11047325 annotated to the *SOCS3* gene, involved in the negative regulation of the JAK-STAT pathway and thought to play a role in modulating the outcome of infections and autoimmune diseases^37^ (effect size per week 0.0008, p = 1.91 x 10^-41^).

**Figure 2:**
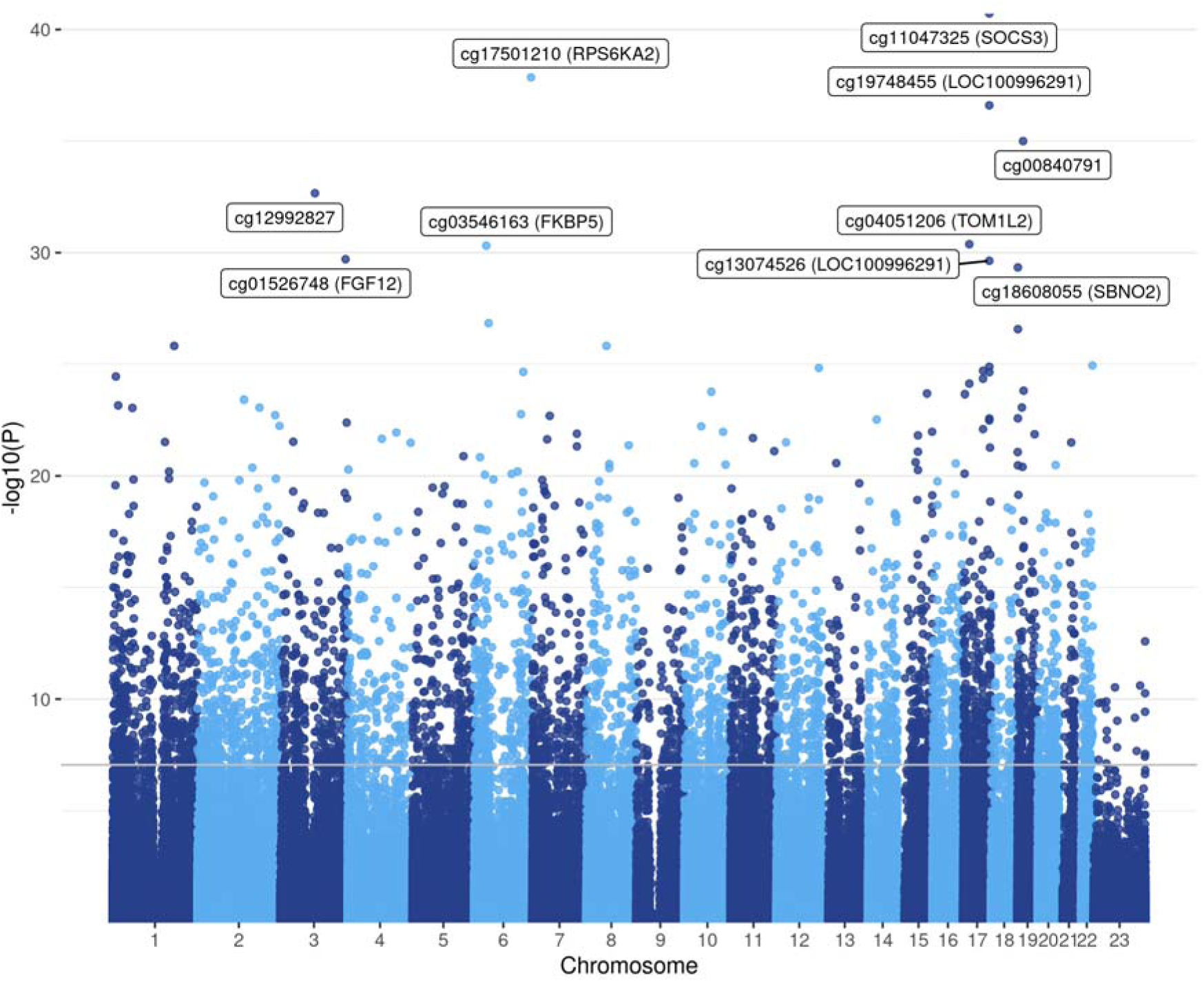
Manhattan plot of CpG sites associated with change over time following anti-TNF drug exposure regardless of treatment outcome. The top 10 differentially methylated probes with annotations are labelled in the plot. The grey horizontal line represents the significant p value threshold of 9 x 10^-8^.

**Table 2:** Top 10 differentially methylated probes associated with anti-TNF treatment over time

| CpG | chr | pos | Relation to Island | Gene name | Gene group | Coefficient (per week) | P value |
| --- | --- | --- | --- | --- | --- | --- | --- |
| cg11047325 | 17 | 76354934 | Island | SOCS3 | Body | 0.0008 | 1.91E-41 |
| cg17501210 | 6 | 166970252 | OpenSea | RPS6KA2 | Body | 0.0007 | 1.40E-38 |
| cg19748455 | 17 | 76274856 | OpenSea | LOC100996291 | TSS1500 | 0.0008 | 2.55E-37 |
| cg00840791 | 19 | 16453259 | OpenSea |  |  | 0.0011 | 1.02E-35 |
| cg12992827 | 3 | 101901234 | OpenSea |  |  | 0.0010 | 2.23E-33 |
| cg04051206 | 17 | 17750855 | OpenSea | TOM1L2 | 3'UTR | 0.0005 | 4.19E-31 |
| cg03546163 | 6 | 35654363 | N_Shore | FKBP5 | 5'UTR | 0.0012 | 4.87E-31 |
| cg01526748 | 3 | 191930926 | OpenSea | FGF12 | Body | -0.0005 | 1.97E-30 |
| cg13074526 | 17 | 76274743 | OpenSea | LOC100996291 | TSS200 | 0.0008 | 2.32E-30 |
| cg18608055 | 19 | 1130866 | OpenSea | SBNO2 | Body | 0.0008 | 4.57E-30 |

Gene ontology (GO) analysis of genes annotated to treatment-associated DMPs identified 108 significant biological pathways (Supplementary Table 3) further implicating the immune response (immune system process (GO: 0002376, FDR < 0.001), immune response (GO: 0006955, FDR < 0.001), and immune system development (GO:0002520, FDR < 0.001)) alongside pathways related to blood cell differentiation (haematopoietic or lymphoid organ development (GO:0048534, FDR < 0.001) and haemopoiesis (GO: 0030097, FDR < 0.001)) (Supplementary Figure 6).

### DNA methylation in infliximab and adalimumab treated patients

Next, we performed an epigenome-wide association study (EWAS) to identify any DMPs associated with anti-TNF treatment type. Overall, there were no significant DMPs at baseline between anti-TNF naïve Crohn’s disease patients who were subsequently treated with infliximab or adalimumab. Irrespective of primary non-response status, we observed 13 DMPs annotated to 9 genes with significantly different trajectories following treatment with infliximab compared to adalimumab. The top-ranked DMPs between treatments included cg03446165 [annotated to *MMP25*] (effect size per week −0.0004, p = 6.78 x 10^-10^), cg12229367 (effect size per week −0.0003, p = 1.54 x 10^-9^) and cg04790662 [annotated to *PAG1*] (effect size per week −0.0005, p = 2.82 x 10^-9^). (Supplementary Table 4)

### DNA methylation differences at baseline are associated with anti-TNF drug concentration following treatment

We sought to determine if DNA methylation difference at baseline prior to the start of anti-TNF treatment were associated with anti-TNF drug concentrations at week 14. We identified 323 DMPs annotated to 210 genes at baseline associated with anti-TNF drug concentrations at week 14 (Table 3, Figure 3). The top ranked DMP was cg23320029 annotated to the *TNIK* gene (effect size 0.0555, p = 4.62 x 10^-15^), encoding the TRAF2 and NCK-interacting kinase, a key regulator in the Wnt signalling pathway implicated in the modulation of immune response during inflammation^38^. GO analysis of genes annotated to DMPs associated with anti-TNF drug concentration at week 14, however, did not identify any FDR (FDR < 0.05) significant pathways.

**Figure 3:**
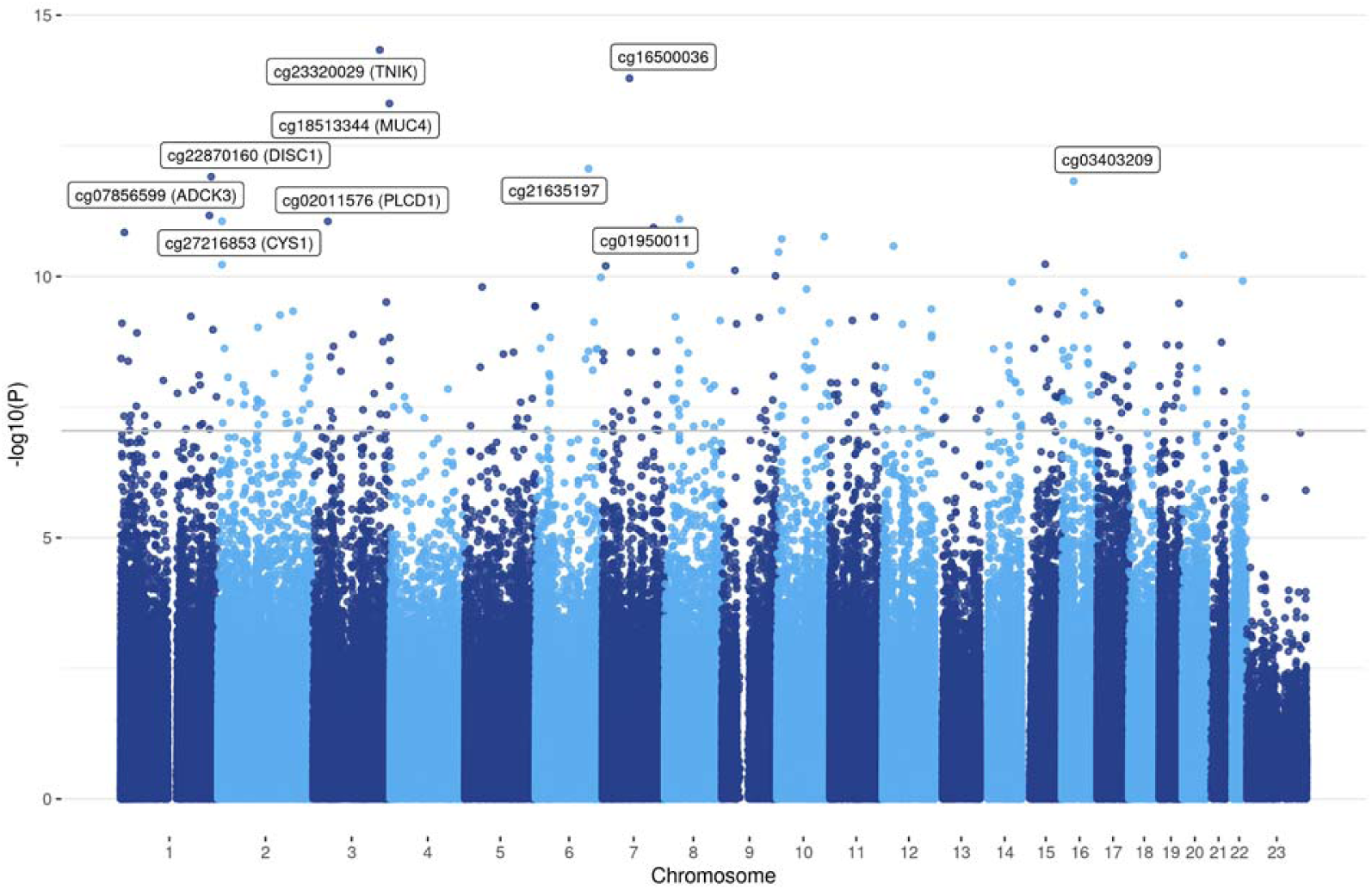
Manhattan plot of CpG sites at baseline associated with anti-TNF drug concentration at week 14. The top 10 CpG sites with their associated gene annotations are labelled in brackets. The grey horizontal line represents the significant p value threshold of 9 x 10^-8^.

**Table 3:** Top 10 differentially methylated probes at baseline associated with anti-TNF drug concentration at week 14

| CpG | chr | pos | Relation to Island | Gene name | Gene group | Coefficient (per week) | P value |
| --- | --- | --- | --- | --- | --- | --- | --- |
| cg23320029 | 3 | 171004750 | OpenSea | TNIK | Body | 0.0555 | 4.62E-15 |
| cg16500036 | 7 | 68983906 | OpenSea |  |  | -0.0409 | 1.61E-14 |
| cg18513344 | 3 | 195531298 | OpenSea | MUC4 | Body | 0.0299 | 4.90E-14 |
| cg21635197 | 6 | 135405808 | OpenSea |  |  | 0.0389 | 8.71E-13 |
| cg22870160 | 1 | 231830793 | OpenSea | DISC1 | Body | -0.0376 | 1.23E-12 |
| cg03403209 | 16 | 29666641 | OpenSea |  |  | -0.0247 | 1.51E-12 |
| cg07856599 | 1 | 227151843 | OpenSea | ADCK3 | Body | -0.0411 | 6.82E-12 |
| cg01950011 | 8 | 37396511 | OpenSea |  |  | -0.0213 | 8.00E-12 |
| cg27216853 | 2 | 10205672 | OpenSea | CYS1 | Body | -0.0371 | 8.79E-12 |
| cg02011576 | 3 | 38060265 | OpenSea | PLCD1 | Body | -0.0387 | 8.82E-12 |

We intersected the list of DMPs predicting anti-TNF drug concentration following treatment with the EWAS catalog^35^ to identify overlaps with DNA methylation differences associated with other traits and diseases, finding that 125 (38.7%) DMPs have been previously associated with other common traits (Supplementary Table 5). The most common shared association (proportion of shared CpGs compared to DMPs) was with an EWAS of body mass index (23.2%), followed by CRP (11.5%), smoking (7.4%), alcohol consumption per day (7.1%) and IBD type (6.8%). The associations with these common traits all had an opposite direction of effect to anti-TNF drug concentration in our cohort; CpG sites associated with a higher BMI and increased CRP were associated with lower anti-TNF drug concentrations, in keeping with the known associations with anti-TNF drug concentration and treatment outcomes^5^.

To understand if there was a relationship between anti-TNF drug concentration at week 14 and anti-TNF treatment response, we performed an EWAS of primary non-response to anti-TNF, and identified 48 DMPs annotated to 36 genes at baseline. Of these, 20 DMPs were associated with both anti-TNF drug concentration and primary non-response to anti-TNF (Supplementary Table 6). These DMPs include cg27216853 [*CYS1*] (effect size to drug concentration −0.0371 vs effect size to primary non-response 0.0245), cg23606775 [*CLSTN1*] (−0.0220 vs 0.0133) and cg18138532 [*UPF2]* (−0.0273 vs 0.0157). Overall, there was a strong correlation of the coefficients (Spearman’s rho = −0.94, p < 0.001) (Figure 4), suggesting a relationship between DMPs associated with lower anti-TNF drug concentration and primary non-response.

**Figure 4:**
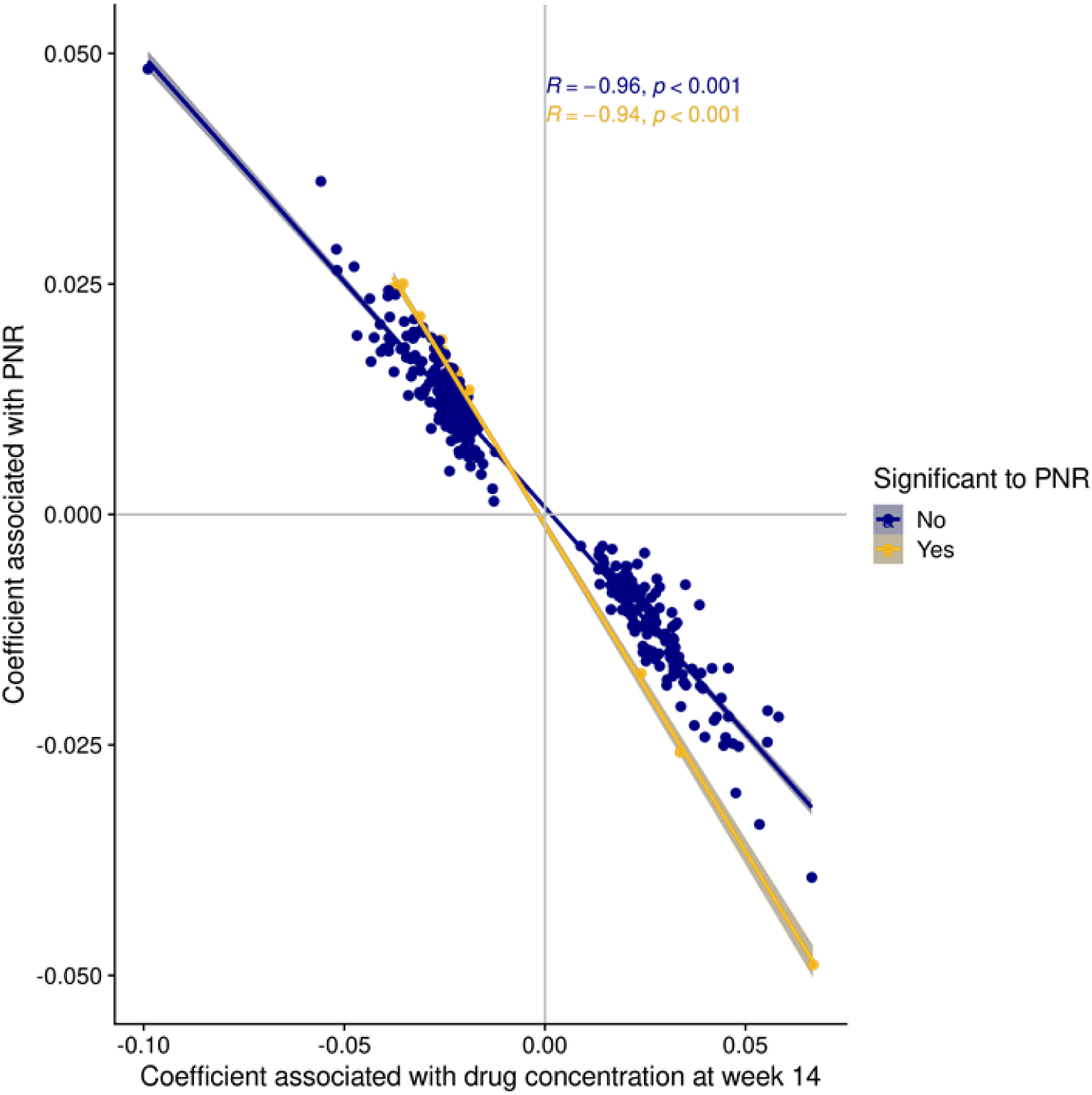
Coefficients of DMPs associated with anti-TNF drug concentration at week 14 and primary non-response. Coefficients represent the beta values of each CpG from linear mixed effects model to each outcome. Spearman’s rho correlation of the coefficients calculated for those that were associated with both anti-TNF drug concentration and primary non-response, and the remaining that were only significant to anti-TNF drug concentration. Abbreviations: PNR = primary non-response.

### Longitudinal changes in DNA methylation differ in patients with primary non-response to anti-TNF treatment

Following anti-TNF treatment, intra-individual changes in DNA methylation was significantly different between those who experienced primary non-response to anti-TNF compared to those who did not at 5 DMPs. These sites were cg07839457 [annotated to *NLRC5*] (effect size per week - 0.0007, p = 1.92 x 10^-13^), cg11047325 [annotated to *SOCS3*] (−0.0007, p = 3.70 x 10^-11^), cg15022400 [annotated to *TRIM69*] (−0.0003, p = 3.39 x 10^-9^), cg25867318 [annotated to *STAT3*] (−0.0004, p = 6.06 x 10^-8^) and cg08950751 [annotated to *AIP*] (−0.0003, p = 6.82 x 10^-8^) (Supplementary Table 7). The top ranked DMP following anti-TNF treatment cg11047325 annotated to *SOCS3* involved in regulation of the JAK-STAT pathway was again identified.

## Discussion

### Key results

In whole blood, we observed almost 5,000 DMPs annotated to >2,000 genes that are associated with anti-TNF therapy, with the genes annotated to these sites being enriched for biological processes related to immune system processes. 323 DMPs annotated to 210 genes were associated with anti-TNF drug concentration at week 14 and we observed an overlap between differentially methylated probes associated with drug concentrations and primary non-response.

### Interpretation

It is perhaps unsurprising, that treatment with the anti-TNF monoclonal antibodies infliximab and adalimumab led to a significant number of differentially methylated probes across multiple genes that were enriched in immune system pathways. Overall, however, only 13 DMPs were found when comparing infliximab- and adalimumab-treated patients, suggesting that there is an anti-TNF treatment class effect and that both drugs exert a similar effect upon levels of DNA methylation. A similar conclusion was made from a study of patients with rheumatoid arthritis treated with several different anti-TNFs including adalimumab, certolizumab, etanercept, golimumab and infliximab, with no DMPs were identified between different anti-TNF subtypes^39^.

The immune cell changes and intracellular signalling pathways in peripheral blood and intestinal tissue following treatment with anti-TNF in patients with IBD is still unclear^40^. Unlike a previous study of 14 patients with IBD^41^, following anti-TNF treatment, we did not observe a change in derived granulocyte proportions between non-responders and responders, but noted differences in B cells and CD4 T cells. Derived cell proportions at baseline were, however, not useful as a biomarker of anti-TNF non-response.

About a third (38.7%) of the DMPs associated with low drug concentrations were linked to other common traits including body mass index, smoking, and CRP that in the PANTS cohort were associated with drug concentration and anti-TNF treatment failure^5^. It is plausible that these DMPs could be used as blood biomarkers independent of clinical traits to predict inter-individual variability in anti-TNF drug concentration, allowing early effective anti-TNF dose prescribing. Our findings that the DMPs were enriched in gene bodies may suggest a more complex mechanism apart from gene transcription in their role underlying anti-TNF treatment response. The role of gene body methylation is still widely debated, and while they have been associated with the regulation of gene expression, they have a more complex role in suppressing aberrant gene transcription and regulating alternative splicing^42^. With the advancement of single-cell sequencing technologies, the study of specific cell types in both disease specific intestinal tissue^43^ and peripheral whole blood^44^perhaps based on our data focussing on the role of B- and CD4^+^ T cells, may provide further insights into the molecular mechanisms underlying anti-TNF treatment failure.

Whilst there was a strong correlation of effect between DMPs associated with lower drug concentration at week 14 and primary non-response, the modest effect sizes mean that these markers are unlikely to be useful as a diagnostic predictor of primary non-response in individual patients. Why primary non-response is so difficult to predict in patients with IBD is unclear. Few of the so-called precision medicine biomarkers to facilitate the right drug, to the right patient, at the right time have been replicated or translated to clinical care. There are a number of possible reasons for this. Firstly, the challenges of defining primary non-response in the absence of endoscopic outcome data. In the PANTS study, we used a pragmatic composite outcome closely linked to routine clinical care that included patient symptoms assessed using a validated severity scores and serum CRP. However, there is poor concordance between symptoms and biomarkers and mucosal inflammation. Patients with Crohn’s disease may also complain of symptoms suggestive of active disease because of overlapping irritable bowel syndrome, bile acid malabsorption and or small intestinal overgrowth. Further interpretation of potential markers across studies is challenging due to differences in study design, inclusion criteria, improvements in experimental and computational methods over time, and critically confounding by sampling of different tissues and cellular heterogeneity. These challenges may explain why we were unable to replicate here the previous associations with oncostatin M (*OSM*)^9, 10^ and triggering receptor expressed on myeloid cells (*TREM-1*)^11, 12^ identified as potential biomarkers predicting non-response to anti-TNF treatment. Our data argues against the presence of a single epigenetic biomarker in whole blood with clinical utility.

Our observations that higher DNA methylation epigenetic smoking score and smoking status, and epigenetic age of participants and chronological age were highly positively correlated internally validates our DNA methylation processing and quality control methods supporting subsequent findings against clinical outcomes. Interestingly, increase in smoking score was observed in all groups regardless of smoking status over time, but was significantly less in former smokers compared to never smokers. Prior longitudinal studies of DNA methylation changes following smoking cessation have reported conflicting results^37, 38^, although varying follow-up times and the study of different populations makes it difficult to compare across studies. Whether the DNA methylation changes following smoking cessation have an impact on anti-TNF drug concentration or outcomes in patients with Crohn’s disease requires further investigation.

### Limitations and generalisability

We acknowledge some important limitations of our work. First, our outcome data could be strengthened with endoscopic outcomes. However, we observed a significant association between clinical outcomes at week 14 and week 54 and faecal calprotectin, which has been shown to closely correlate with endoscopic findings. Second, we measured DNA methylation from whole blood which is likely to be confounded by differences in individual cell proportions. Although we included derived cell proportions as a covariate in our statistical models, this is unlikely to fully control for cellular changes which may be better controlled for by expanded panels of blood cell types or single-cell analyses. Whether similar changes also occur in the target tissues in the small and large intestine is unknown. Third, our findings should be validated in an independent cohort prior to translation into clinical practice.

The PANTS study recruited patients from across the UK, and we believe our findings will be generalisable to patients with Crohn’s disease treated with an anti-TNF across other western populations. Further work is required to determine if these findings are found in other non-western populations, and indeed in other populations of patients with IBD such as those with ulcerative colitis, and in non-IBD patients treated with an anti-TNF.

## Conclusion

Baseline DNA methylation profiles may be used as a predictor for anti-TNF drug concentration at week 14 to identify patients who may benefit from dose optimisation at the outset of anti-TNF therapy.

## Data Availability

Individual participant de-identified data that underlie the results reported in this article will be available immediately after publication for a period of 5 years. The data will be made available to investigators whose proposed use of the data has been approved by an independent review committee. Analyses will be restricted to the aims in the approved proposal. Proposals should be directed to. To gain access data requestors will need to sign a data access agreement.

## Acknowledgements

S.L. is supported by a Wellcome GW4-CAT fellowship (222850/Z/21/Z). N.C. acknowledges support from Crohn’s and Colitis UK. This study was supported by the National Institute for Health and Care Research (NIHR) Exeter Biomedical Research Centre. The views expressed are those of the author(s) and not necessarily those of the NIHR or the Department of Health and Social Care. The authors would like to acknowledge James Butler for facilitating this collaboration and Samantha Lent, Justin Wade Davis, Elina Regan, Stephen Abel, Elizabeth Asque and Areej Ammar in the preparation of this manuscript. We also acknowledge John Kirkwood and Anna Barnes from the NIHR Clinical Research Facility for their support with sample preparation, and the study coordinators of the Exeter IBD Research Group, Marian Parkinson and Helen Gardner-Thorpe for their ongoing administrative support to the study.

## Author Contributions

NAK, JRG, TA participated in the conception and design of this study. CB was the project manager and coordinated patient recruitment. SL, EH, NS, JFW, VP, HG, CB, NC, BB, NAK, JRG, JM, TA were involved in the acquisition, analysis or interpretation of data. Data analysis was done by SL and EH. Drafting of the manuscript was done by SL, EH, NS, JFW, VP, HG, NC, BB, NAK, JRG, JM, TA. TA obtained the funding for the study. All the authors contributed to the critical review and final approval of the manuscript. JM and TA have verified the underlying data.

