## Supplementary information for "Whole blood DNA methylation changes are associated with anti-TNF drug concentration in patients with Crohn’s disease"

#### Supplementary Table 1: IBD Pharmacogenetics Study Group

All UK gastroenterologists were invited to participate in the PANTS study which was promoted through the UK National Institute for Health Research and the British Society of Gastroenterology.

| Hospital or Trust name | City | Name | Job Title |
|---|---|---|---|
| Tameside Hospital NHS Foundation Trust | Ashton U Lyne | Dr Vinod Patel | Consultant Gastroenterologist |
| Basildon and Thurrock University Hospitals NHS Foundation Trust | Basildon | Dr Zia Mazhar | Consultant Gastroenterologist |
| Hampshire Hospitals NHS Foundation Trust | Basingstoke | Dr Rebecca Saich | Consultant Gastroenterologist |
| Royal United Hospital | Bath | Dr Ben Colleypriest | Consultant Gastroenterologist |
| Ulster Hospital | Belfast | Dr Tony C Tham | Consultant Gastroenterologist |
| University Hospital's Birmingham NHS Foundation Trust | Birmingham | Dr Tariq H Iqbal | Consultant Gastroenterologist |
| East Lancashire NHS Teaching Trust | Blackburn | Dr Vishal Kaushik | Consultant Gastroenterologist |
| Blackpool Teaching Hospitals NHS Foundation Trust | Blackpool | Dr Senthil Murugesan | Consultant Gastroenterologist |
| Bolton NHS Trust | Bolton | Dr Salil Singh | Consultant Gastroenterologist |
| Royal Bournemouth Hospital | Bournemouth | Dr Sean Weaver | Consultant Gastroenterologist |
| Bradford Teaching Hospitals Foundation Trust - (St Lukes Hospital &Bradford Royal Infirmary) | Bradford | Dr Cathryn Preston | Consultant Gastroenterologist |
| Brighton and Sussex University Hospitals NHS Trust | Brighton | Dr Assad Butt | Paediatric Consultant Gastroenterologist |
| Brighton and Sussex University Hospitals NHS Trust | Brighton | Dr Melissa Smith | Consultant Gastroenterologist |
| University Hospitals Bristol NHS Foundation Trust | Bristol | Dr Dharamveer Basude | Consultant Paediatric Gastroenterologist |
| University Hospitals Bristol NHS Foundation Trust | Bristol | Dr Amanda Beale | Consultant Gastroenterologist |

| | | | |
|---|---|---|---|
| Frimley Park Hospital NHS Foundation Trust | Camberley | Dr Sarah Langlands | Consultant Gastroenterologist |
| Frimley Park Hospital NHS Foundation Trust | Camberley | Dr Natalie Direkze | Consultant gastroenterologist |
| Cambridge University Hospitals NHS Foundation Trust | Cambridge | Dr Miles Parkes | Consultant Gastroenterologist |
| Cambridge University Hospitals NHS Foundation Trust | Cambridge | Dr Franco Torrente | Consultant Paediatric Gastroenterologist |
| Cambridge University Hospitals NHS Foundation Trust | Cambridge | Dr Juan De La Revella Negro | Research fellow |
| North Cumbria University Hospitals NHS Trust | Carlisle | Dr Chris Ewen MacDonald | Consultant Gastroenterologist |
| Ashford & St Peter's Hospitals NHS Foundation Trust | Chertsey | Dr Stephen M Evans | Consultant Gastroenterologist |
| St Peter's Hospital | Chertsey | Dr Anton V J Gunasekera | Consultant Gastroenterologist |
| Ashford & St Peter's Hospitals NHS Foundation Trust | Chertsey | Dr Alka Thakur | Paediatric Consultant |
| Chesterfield Royal NHS Foundation Trust | Chesterfield | Dr David Elphick | Consultant Gastroenterologist |
| Colchester Hospital University NHS Foundation Trust | Colchester | Dr Achuth Shenoy | Consultant Gastroenterologist |
| University Hospitals Coventry and Warwickshire NHS Trust | Coventry | Prof Chuka U Nwokolo | Consultant Gastroenterologist |
| County Durham and Darlington NHS Foundation Trust | Darlington | Dr Anjan Dhar | Consultant Gastroenterologist & Hon. Clinical Lecturer |
| Derby Hospital NHS Foundation NHS Trust | Derby | Dr Andrew T Cole | Consultant Gastroenterologist |
| Doncaster and Bassetlaw Hospitals NHS Foundation Trust | Doncaster | Dr Anurag Agrawal | Consultant Gastroenterologist |
| Dorset County Hospital NHS Foundation Trust | Dorchester | Dr Stephen Bridger | Consultant Gastroenterologist |
| Dorset County Hospitals Foundation Trust | Dorchester | Dr Julie Doherty | Paediatric Consultant |

| | | | |
|---|---|---|---|
| Dudley Group NHS Foundation Trust | Dudley | Dr Sheldon C Cooper | Consultant Gastroenterologist |
| Russells Hall Hospital, The Dudley Group NHS Foundation Trust | Dudley | Dr Shanika de Silva | Consultant Gastroenterologist |
| Ninewells Hospital & Medical School | Dundee | Dr Craig Mowat | Consultant Gastroenterologist |
| East Sussex Healthcare Trust | Eastborne | Dr Phillip Mayhead | Consultant Gastroenterologist |
| NHS Lothian | Edinburgh | Dr Charlie Lees | Consultant Gastroenterologist and Honorary Senior Lecturer |
| NHS Lothian | Edinburgh | Dr Gareth Jones | Research fellow |
| Royal Devon and Exeter NHS Foundation Trust | Exeter | Dr Tariq Ahmad | Consultant Gastroenterologist |
| Royal Devon and Exeter NHS Foundation Trust | Exeter | Dr James W Hart | Consultant Paediatrician |
| Glasgow Royal Infirmary | Glasgow | Dr Daniel R Gaya | Consultant Gastroenterologist |
| Royal Hospital for Children | Glasgow | Prof Richard K Russell | Consultant Paediatric Gastroenterologist |
| Royal Hospital for Children | Glasgow | Dr Lisa Gervais | Research fellow |
| Gloucestershire Hospitals NHS Trust | Gloucester | Dr Paul Dunckley | Consultant Gastroneterologist |
| United Lincolnshire Hospitals NHS Trust | Grantham | Dr Tariq Mahmood | Consultant Gastroenterologist |
| James Paget University Hospitals NHS Foundation Trust | Great Yarmouth | Dr Paul J R Banim | Consultant Gastroneterologist |
| Calderdale and Huddersfield NHS Trust | Halifax | Dr Sunil Sonwalkar | Consultant Gastroenterologist |
| Princess Alexandra Hospital NHS Trust | Harlow | Dr Deb Ghosh | Consultant Gastroenterologist |
| Princess Alexandra Hospital NHS Trust | Harlow | Dr Rosemary H Phillips | Consultant Gastroenterologist |
| Hull and East Yorkshire NHS Trust | Hull | Dr Amer Azaz | Paediatric Consultant Gastroenterologist |

| | | | |
|---|---|---|---|
| Hull and East Yorkshire NHS Trust | Hull | Dr Shaji Sebastian | Consultant Gastroenterologist |
| Airedale NHS Foundation Trust | Keighley | Dr Richard Shenderey | Consultant Gastroenterologist |
| Crosshouse Hospital | Kilmarnock | Dr Lawrence Armstrong | Consultant Paediatrician |
| Crosshouse Hospital | Kilmarnock | Dr Claire Bell | Research fellow |
| The Queen Elizabeth Hospital NHS Foundation Trust | Kings Lynn | Dr Radhakrishnan Hariraj | Consultant Gastroenterologist |
| Kingston Hospital NHS Trust | Kingston upon Thames | Dr Helen Matthews | Consultant Gastroenterologist |
| NHS Fife | Kirkcaldy | Dr Hasnain Jafferbhoy | Consultant Gastroenterologist |
| Leeds Teaching Hospitals NHS Trust | Leeds | Dr Christian P Selinger | Consultant Gastroenterologist |
| Leeds Teaching Hospitals NHS Trust | Leeds | Dr Veena Zamvar | Paediatric Consultant Gastroenteorlogist |
| University Hospitals of Leicester NHS Trust | Leicester | Prof John S De Caestecker | Consultant Gastroenterologist |
| University Hospitals of Leicester NHS Trust | Leicester | Dr Anne Willmott | Paediatric Consultant Gastroenterologist |
| Mid Cheshire Hospitals NHS Foundation Trust | Leighton | Mr Richard Miller | Research Nurse |
| United Lincolnshire Hospitals NHS Trust | Lincoln | Dr Palani Sathish Babu | Consultant Gastroenterologist |
| Alder Hey Childrens Hospital | Liverpool | Dr Christos Tzivinikos | Consultant Paediatric Gastroenterologist |
| University College London Hospitals NHS Foundation Trust | London | Dr Stuart L Bloom | Consultant Gastroenterologist |
| Kings College Hospital NHS Foundation Trust | London | Dr Guy Chung-Faye | Consultant Gastroenterologist |
| Royal London Childrens Hospital, Barts Health NHS Trust | London | Prof Nicholas M Croft | Paediatric Consultant Gastroenterologist |
| Chelsea & Westminster Hospital | London | Dr John ME Fell | Consultant Paediatric Gastroenterologist |

| | | | |
|---|---|---|---|
| Chelsea and Westminster Hospital NHS Foundation | London | Dr Marcus Harbord | Consultant Gastroenterologist |
| North West London Hospitals NHS Trust | London | Dr Ailsa Hart | Consultant Gastroenterologist |
| Kings College Hospital NHS Foundation Trust | London | Dr Ben Hope | Consultant Paediatrician |
| Guys & St Thomas' NHS Foundation Trust | London | Dr Peter M Irving | Consultant Gastroenterologist |
| Barts and The London NHS Trust | London | Prof James O Lindsay | Consultant Gastroenterologist |
| Guy's and St Thomas' NHS trust | London | Dr Joel E Mawdsley | Gastroenterology Consultant |
| Lewisham and Greenwich Healthcare NHS Trust | London | Dr Alistair McNair | Consultant Gastroenterologist |
| Chelsea and Westminster Hospital NHS Foundation | London | Dr Kevin J Monahan | Consultant Gastroenterologist |
| Royal Free London NHS Foundation Trust | London | Dr Charles D Murray | Consultant Gastroenterologist |
| Imperial College Healthcare NHS Trust | London | Prof Timothy Orchard | Consultant Gastroenterologist |
| St George's Healthcare NHS Trust | London | Dr Thankam Paul | Paediatric Consultant Gastroenterologist |
| St George's Healthcare NHS Trust | London | Dr Richard Pollok | Reader and Consultant Gastroenterologist |
| Great Ormond Street Hospital for Children NHS Foundation Trust | London | Dr Neil Shah | Consultant Gastroenterologist |
| North West London Hospitals NHS Trust | London | Dr Sonia Bouri | Research fellow |
| The Luton & Dunstable University Hospital | Luton | Dr Matt W Johnson | Consultant Gastroenterologist |
| Luton and Dunstable Hospital Foundation Trust | Luton | Dr Anita Modi | Paediatric Consultant with Allergy and Gastroenterology interest |
| The Luton & Dunstable University Hospital | Luton | Dr Kasamu Dawa Kabiru | Research fellow |

| | | | |
|---|---|---|---|
| Maidstone and Tunbridge Wells NHS Trust | Maidstone | Dr B K Baburajan | Consultant Gastroenterologist |
| Maidstone and Tunbridge Wells NHS Trust | Maidstone | Prof Bim Bhaduri | Paediatric Consultant Gastroenterologist |
| Manchester University Hospitals NHS Foundation Trust | Manchester | Dr Andrew Adebayo Fagbemi | Consultant Gastroenterologist |
| Central Manchester University Hospitals NHS Foundation Trust | Manchester | Dr Scott Levison | Consultant Gastroenterologist |
| The Pennine Acute Hospitals NHS Trust | Manchester | Dr Jimmy K Limdi | Consultant Gastroenterologist |
| Manchester University NHS Foundation Trust, Wythenshawe Hospital | Manchester | Dr Gill Watts | Consultant Gastroenterologist |
| Sherwood Forest Hospitals NHS Foundation Trust | Mansfield | Dr Stephen Foley | Consultant Gastroenterologist |
| South Tees Hospital NHS Foundation Trust | Middlesbrough | Dr Arvind Ramadas | Consultant Gastroenterologist |
| Milton Keynes Hospital NHS Foundation Trust | Milton Keynes | Dr George MacFaul | Consultant Gastroenterologist |
| Newcastle Upon Tyne Hospital Trust | Newcastle | Dr John Mansfield | Consultant Gastroenterologist |
| Isle of Wight NHS Foundation Trust | Newport | Dr Leonie Grellier | Consultant Gastroenterologist |
| Norfolk & Norwich University Hospital NHS Foundation Trust | Norwich | Dr Mary-Anne Morris | Consultant Paediatric Gastroenterologist |
| Norfolk & Norwich University Hospital NHS Foundation Trust | Norwich | Dr Mark Tremelling | Consultant Gastroenterologist |
| Nottingham University Hospitals NHS Trust | Nottingham | Prof Chris Hawkey | Consultant Gastroenterologist |
| Nottingham University Hospitals NHS Trust | Nottingham | Dr Sian Kirkham | Consultant Paediatric Gastroenterologist |
| Nottingham University Hospitals NHS Trust | Nottingham | Dr Charles PJ Charlton | Consultant gastroenterologist |
| Oxford University Hospitals NHS Foundation Trust | Oxford | Dr Astor Rodrigues | Paediatric Consultant Gastroenterologist |

| | | | |
|---|---|---|---|
| Oxford University Hospitals NHS Trust | Oxford | Prof Alison Simmons | Consultant Gastroenterologist |
| Plymouth Hospitals NHS Trust | Plymouth | Dr Stephen J Lewis | Consultant Gastroenterologist |
| Poole Hospital NHS Foundation Trust | Poole | Dr Jonathon Snook | Consultant Gastroenterologist |
| Poole Hospital NHS Foundation Trust | Poole | Dr Mark Tighe | Paediatric Consultant with interest in Oncology and Gastroenterology |
| Portsmouth Hospitals NHS Trust | Portsmouth | Dr Patrick M Goggin | Consultant Gastroenterologist |
| Royal Berkshire NHS Foundation Trust | Reading | Dr Aminda N De Silva | Consultant Gastroenterologist |
| Salford Royal NHS Foundation Trust | Salford | Prof Simon Lal | Consultant Gastroenterologist |
| Shrewsbury and Telford Hospital NHS Trust | Shrewsbury | Dr Mark S Smith | Consultant Gastroenterologist |
| South Tyneside NHS Foundation Trust | South Shields | Dr Simon Panter | Consultant Gastroenterologist |
| Southampton University Hospitals NHS Trust | Southampton | Dr Fraser Cummings | Consultant Gastroenterologist |
| Southampton University Hospitals NHS Trust | Southampton | Dr Suranga Dharmisari | Research fellow |
| East and North Herts NHS Trust | Stevenage | Dr Martyn Carter | Consultant Gastroenterologist |
| NHS Forth Valley | Stirling | Dr David Watts | Consultant Gastroenterologist |
| Stockport NHS foundation Trust | Stockport | Dr Zahid Mahmood | Consultant Gastroenterologist |
| North Tees and Hartlepool NHS Foundation Trust | Stockton | Dr Bruce McLain | Paediatric Consultant Gastroenterologist |
| University Hospitals of North Staffordshire | Stoke-on Trent | Dr Sandip Sen | Consultant Gastroenterologist |
| University Hospitals of North Midlands NHS Trust | Stoke-on-Trent | Dr Anna J Pigott | Consultant Paediatric Gastroenterologist |
| City Hospitals Sunderland NHS Foundation Trust | Sunderland | Dr David Hobday | Consultant Gastroenterologist |

| | | | |
|---|---|---|---|
| Taunton and Somerset NHS Foundation Trust | Taunton | Dr Emma Wesley | Consultant Gastroenterologist |
| South Devon Healthcare NHS Foundation Trust | Torquay | Dr Richard Johnston | Consultant Gastroenterologist |
| South Devon Healthcare NHS Foundation Trust | Torquay | Dr Cathryn Edwards | Consultant gastroenterologist |
| Royal Cornwall Hospitals NHS Trust | Truro | Dr John Beckly | Consultant Gastroenterologist |
| Mid Yorkshire Hospitals NHS Trust | Wakefield | Dr Deven Vani | Consultant Physician & Gastroenterologist |
| Warrington& Halton NHS Foundation | Warrington | Dr Subramaniam Ramakrishnan | Consultant Gastroenterologist |
| West Hertfordshire Hospitals NHS Trust | Watford | Dr Rakesh Chaudhary | Consultant Gastroenterologist |
| Sandwell and West Birmingham Hospitals NHS Trust | West Bromwich | Dr Nigel J Trudgill | Consultant Gastroenterologist |
| Sandwell and West Birmingham Hospitals NHS Trust | West Bromwich | Dr Rachel Cooney | Consultant gastroenterologist |
| Weston Area Health NHS Trust | Weston-Super-Mare | Dr Andy Bell | Consultant Gastroenterologist |
| Royal Albert Edward Infirmary, Wrightington, Wigan & Leigh NHS Foundation Trust | Wigan | Dr Neeraj Prasad | Consultant Gastroenterologist |
| Hampshire Hospitals NHS Foundation Trust | Winchester | Dr John N Gordon | Consultant Gastroenterologist |
| Royal Wolverhampton Hospitals NHS Trust | Wolverhampton | Prof Matthew J Brookes | Consultant Gastroenterologist |
| Western Sussex Hospitals NHS Trust | Worthing | Dr Andy Li | Consultant Gastroenterologist |
| Yeovil District Hospital NHS Foundation Trust | Yeovil | Dr Stephen Gore | Consultant Gastroenterologist |

#### Supplementary Table 2: Distribution of samples by individual participant and study visits

| **Distribution of samples by individual participant** | |
|---|---|
| **Number of samples** | **Number of participants** |
| 1 | 34 |
| 2 | 107 |
| 3 | 157 |
| 4 | 87 |
| **Distribution of samples by study visits** | |
| **Study visit (weeks since anti-TNF treatment commenced)** | **Number of samples** |
| Week 0 | 373 |
| Week 14 | 314 |
| Week 30 | 251 |
| Week 54 | 124 |

#### Supplementary Table 3: Gene ontology (GO) terms of differentially methylated probes associated with anti-TNF treatment

| Pathway ID | ONTOLOGY | TERM | N | DE | P.DE | FDR |
|---|---|---|---|---|---|---|
| GO:0002376 | Biological process | immune system process | 2450 | 408 | 2.97E-13 | 6.79E-09 |
| GO:0006955 | Biological process | immune response | 1646 | 267 | 1.28E-11 | 1.47E-07 |
| GO:0002520 | Biological process | immune system development | 914 | 184 | 7.91E-09 | 5.82E-05 |
| GO:0048534 | Biological process | hematopoietic or lymphoid organ development | 859 | 175 | 1.02E-08 | 5.82E-05 |
| GO:0030097 | Biological process | hemopoiesis | 820 | 167 | 1.65E-08 | 7.52E-05 |
| GO:0002682 | Biological process | regulation of immune system process | 1328 | 224 | 2.59E-08 | 9.87E-05 |
| GO:0009607 | Biological process | response to biotic stimulus | 1368 | 207 | 5.80E-08 | 0.000189 |
| GO:0002684 | Biological process | positive regulation of immune system process | 814 | 145 | 1.12E-07 | 0.000319 |
| GO:0006952 | Biological process | defense response | 1579 | 228 | 1.43E-07 | 0.000363 |
| GO:0050776 | Biological process | regulation of immune response | 804 | 141 | 1.83E-07 | 0.000419 |
| GO:0002521 | Biological process | leukocyte differentiation | 536 | 112 | 2.80E-07 | 0.000581 |
| GO:0043207 | Biological process | response to external biotic stimulus | 1329 | 196 | 3.92E-07 | 0.000688 |
| GO:0051707 | Biological process | response to other organism | 1328 | 196 | 3.79E-07 | 0.000688 |
| GO:0001816 | Biological process | cytokine production | 802 | 135 | 4.42E-07 | 0.000722 |
| GO:0001817 | Biological process | regulation of cytokine production | 789 | 132 | 7.71E-07 | 0.001175 |
| GO:0045321 | Biological process | leukocyte activation | 841 | 151 | 1.35E-06 | 0.001926 |
| GO:0001819 | Biological process | positive regulation of cytokine production | 463 | 89 | 1.78E-06 | 0.002154 |
| GO:0098542 | Biological process | defense response to other organism | 972 | 143 | 1.79E-06 | 0.002154 |
| GO:1903131 | Biological process | mononuclear cell differentiation | 420 | 90 | 1.77E-06 | 0.002154 |
| GO:0044419 | Biological process | biological process involved in interspecies interaction between organisms | 1480 | 218 | 2.01E-06 | 0.002297 |
| GO:0030217 | Biological process | T cell differentiation | 256 | 62 | 2.46E-06 | 0.002677 |
| GO:0043367 | Biological process | CD4-positive, alpha-beta T cell differentiation | 83 | 28 | 2.94E-06 | 0.003049 |
| GO:0045087 | Biological process | innate immune response | 753 | 116 | 3.44E-06 | 0.003419 |

| | | | | | | |
|---|---|---|---|---|---|---|
| GO:0001775 | Biological process | cell activation | 978 | 172 | 3.89E-06 | 0.003704 |
| GO:0030098 | Biological process | lymphocyte differentiation | 368 | 80 | 4.07E-06 | 0.003721 |
| GO:0030155 | Biological process | regulation of cell adhesion | 743 | 155 | 4.45E-06 | 0.003906 |
| GO:0070665 | Biological process | positive regulation of leukocyte proliferation | 150 | 37 | 5.82E-06 | 0.00492 |
| GO:0005085 | Biological process | guanyl-nucleotide exchange factor activity | 223 | 68 | 6.19E-06 | 0.005053 |
| GO:0042110 | Biological process | T cell activation | 478 | 94 | 7.67E-06 | 0.006044 |
| GO:0007159 | Biological process | leukocyte cell-cell adhesion | 366 | 73 | 1.02E-05 | 0.007511 |
| GO:0071674 | Biological process | mononuclear cell migration | 191 | 43 | 1.02E-05 | 0.007511 |
| GO:0006909 | Biological process | phagocytosis | 232 | 58 | 1.12E-05 | 0.007805 |
| GO:0070663 | Biological process | regulation of leukocyte proliferation | 242 | 51 | 1.13E-05 | 0.007805 |
| GO:0046632 | Biological process | alpha-beta T cell differentiation | 112 | 34 | 1.20E-05 | 0.008083 |
| GO:0046649 | Biological process | lymphocyte activation | 685 | 126 | 1.25E-05 | 0.008145 |
| GO:0070661 | Biological process | leukocyte proliferation | 312 | 61 | 1.46E-05 | 0.009287 |
| GO:1903706 | Biological process | regulation of hemopoiesis | 362 | 76 | 1.84E-05 | 0.011378 |
| GO:0050900 | Biological process | leukocyte migration | 363 | 71 | 2.75E-05 | 0.016531 |
| GO:0002252 | Biological process | immune effector process | 570 | 98 | 3.28E-05 | 0.017396 |
| GO:0006954 | Biological process | inflammatory response | 793 | 123 | 3.35E-05 | 0.017396 |
| GO:0032609 | Biological process | interferon-gamma production | 110 | 26 | 3.20E-05 | 0.017396 |
| GO:0032649 | Biological process | regulation of interferon-gamma production | 110 | 26 | 3.20E-05 | 0.017396 |
| GO:0035710 | Biological process | CD4-positive, alpha-beta T cell activation | 100 | 29 | 3.03E-05 | 0.017396 |
| GO:0051246 | Biological process | regulation of protein metabolic process | 2536 | 395 | 3.35E-05 | 0.017396 |
| GO:1903039 | Biological process | positive regulation of leukocyte cell-cell adhesion | 236 | 52 | 3.69E-05 | 0.018709 |
| GO:0002764 | Biological process | immune response-regulating signaling pathway | 388 | 79 | 3.88E-05 | 0.019256 |
| GO:0002694 | Biological process | regulation of leukocyte activation | 523 | 95 | 4.19E-05 | 0.020129 |
| GO:0050670 | Biological process | regulation of lymphocyte proliferation | 222 | 46 | 4.30E-05 | 0.020129 |
| GO:1903037 | Biological process | regulation of leukocyte cell-cell adhesion | 331 | 65 | 4.32E-05 | 0.020129 |

| | | | | | | |
|---|---|---|---|---|---|---|
| GO:0050671 | Biological process | positive regulation of lymphocyte proliferation | 137 | 32 | 4.41E-05 | 0.020136 |
| GO:0042093 | Biological process | T-helper cell differentiation | 66 | 22 | 4.67E-05 | 0.020928 |
| GO:0032944 | Biological process | regulation of mononuclear cell proliferation | 224 | 46 | 5.02E-05 | 0.021549 |
| GO:0032946 | Biological process | positive regulation of mononuclear cell proliferation | 138 | 32 | 5.01E-05 | 0.021549 |
| GO:0042102 | Biological process | positive regulation of T cell proliferation | 101 | 25 | 5.19E-05 | 0.021549 |
| GO:0048583 | Biological process | regulation of response to stimulus | 3772 | 591 | 5.13E-05 | 0.021549 |
| GO:0002253 | Biological process | activation of immune response | 296 | 61 | 5.41E-05 | 0.022065 |
| GO:0002294 | Biological process | CD4-positive, alpha-beta T cell differentiation involved in immune response | 68 | 22 | 5.75E-05 | 0.022416 |
| GO:0007166 | Biological process | cell surface receptor signaling pathway | 2606 | 434 | 5.61E-05 | 0.022416 |
| GO:0022409 | Biological process | positive regulation of cell-cell adhesion | 281 | 61 | 5.79E-05 | 0.022416 |
| GO:0051249 | Biological process | regulation of lymphocyte activation | 434 | 82 | 5.97E-05 | 0.022717 |
| GO:1902105 | Biological process | regulation of leukocyte differentiation | 277 | 60 | 6.48E-05 | 0.024254 |
| GO:0045785 | Biological process | positive regulation of cell adhesion | 433 | 93 | 6.60E-05 | 0.024332 |
| GO:0002287 | Biological process | alpha-beta T cell activation involved in immune response | 69 | 22 | 6.87E-05 | 0.024504 |
| GO:0002293 | Biological process | alpha-beta T cell differentiation involved in immune response | 69 | 22 | 6.87E-05 | 0.024504 |
| GO:0050870 | Biological process | positive regulation of T cell activation | 213 | 47 | 7.10E-05 | 0.024584 |
| GO:1901701 | Biological process | cellular response to oxygen-containing compound | 1195 | 209 | 7.10E-05 | 0.024584 |
| GO:0050778 | Biological process | positive regulation of immune response | 495 | 86 | 7.41E-05 | 0.024875 |
| GO:0050863 | Biological process | regulation of T cell activation | 324 | 64 | 7.37E-05 | 0.024875 |
| GO:0071216 | Biological process | cellular response to biotic stimulus | 241 | 47 | 7.92E-05 | 0.026226 |
| GO:0051345 | Biological process | positive regulation of hydrolase activity | 581 | 116 | 8.29E-05 | 0.02705 |
| GO:0051251 | Biological process | positive regulation of lymphocyte activation | 284 | 59 | 8.47E-05 | 0.027243 |
| GO:0030695 | Biological process | GTPase regulator activity | 475 | 115 | 9.28E-05 | 0.029052 |
| GO:0060589 | Biological process | nucleoside-triphosphatase regulator activity | 475 | 115 | 9.28E-05 | 0.029052 |
| GO:0045619 | Biological process | regulation of lymphocyte differentiation | 173 | 42 | 9.46E-05 | 0.029212 |
| GO:0002366 | Biological process | leukocyte activation involved in immune response | 268 | 54 | 9.71E-05 | 0.029559 |

| | | | | | | |
|---|---|---|---|---|---|---|
| GO:0030099 | Biological process | myeloid cell differentiation | 378 | 80 | 9.96E-05 | 0.029932 |
| GO:0019221 | Biological process | cytokine-mediated signaling pathway | 461 | 76 | 0.000118 | 0.033198 |
| GO:0043547 | Biological process | positive regulation of GTPase activity | 247 | 64 | 0.000113 | 0.033198 |
| GO:0046637 | Biological process | regulation of alpha-beta T cell differentiation | 68 | 22 | 0.000117 | 0.033198 |
| GO:0046651 | Biological process | lymphocyte proliferation | 282 | 53 | 0.000117 | 0.033198 |
| GO:0050790 | Biological process | regulation of catalytic activity | 2365 | 390 | 0.000116 | 0.033198 |
| GO:0005096 | Biological process | GTPase activator activity | 449 | 110 | 0.000124 | 0.034543 |
| GO:0030595 | Biological process | leukocyte chemotaxis | 226 | 45 | 0.000126 | 0.034611 |
| GO:0030335 | Biological process | positive regulation of cell migration | 550 | 110 | 0.000131 | 0.03532 |
| GO:0043370 | Biological process | regulation of CD4-positive, alpha-beta T cell differentiation | 51 | 17 | 0.00013 | 0.03532 |
| GO:0002292 | Biological process | T cell differentiation involved in immune response | 75 | 22 | 0.000137 | 0.036472 |
| GO:0032943 | Biological process | mononuclear cell proliferation | 285 | 53 | 0.000146 | 0.038322 |
| GO:0002696 | Biological process | positive regulation of leukocyte activation | 329 | 64 | 0.000155 | 0.038876 |
| GO:0006793 | Biological process | phosphorus metabolic process | 2705 | 440 | 0.000152 | 0.038876 |
| GO:0043087 | Biological process | regulation of GTPase activity | 338 | 84 | 0.000151 | 0.038876 |
| GO:0048584 | Biological process | positive regulation of response to stimulus | 2035 | 324 | 0.000153 | 0.038876 |
| GO:0032268 | Biological process | regulation of cellular protein metabolic process | 2367 | 363 | 0.00016 | 0.039822 |
| GO:0050865 | Biological process | regulation of cell activation | 567 | 100 | 0.000174 | 0.042177 |
| GO:0071310 | Biological process | cellular response to organic substance | 2389 | 378 | 0.000174 | 0.042177 |
| GO:0097529 | Biological process | myeloid leukocyte migration | 218 | 43 | 0.000178 | 0.042693 |
| GO:1903749 | Biological process | positive regulation of establishment of protein localization to mitochondrion | 36 | 14 | 0.000181 | 0.043044 |
| GO:0044093 | Biological process | positive regulation of molecular function | 1478 | 262 | 0.000184 | 0.043403 |
| GO:0006796 | Biological process | phosphate-containing compound metabolic process | 2684 | 436 | 0.000187 | 0.043498 |
| GO:1903747 | Biological process | regulation of establishment of protein localization to mitochondrion | 50 | 17 | 0.000189 | 0.043498 |
| GO:0002263 | Biological process | cell activation involved in immune response | 272 | 54 | 0.000197 | 0.044588 |

| | | | | | | |
|---|---|---|---|---|---|---|
| GO:0002274 | Biological process | myeloid leukocyte activation | 220 | 45 | 0.000196 | 0.044588 |
| GO:0050867 | Biological process | positive regulation of cell activation | 340 | 65 | 0.000204 | 0.045585 |
| GO:0051336 | Biological process | regulation of hydrolase activity | 1023 | 178 | 0.000209 | 0.046295 |
| GO:0010033 | Biological process | response to organic substance | 3041 | 458 | 0.000214 | 0.04693 |
| GO:0007259 | Biological process | receptor signaling pathway via JAK-STAT | 166 | 34 | 0.000218 | 0.047393 |
| GO:0072676 | Biological process | lymphocyte migration | 112 | 25 | 0.00022 | 0.047393 |
| GO:0002250 | Biological process | adaptive immune response | 423 | 72 | 0.000223 | 0.047621 |
| GO:0031663 | Biological process | lipopolysaccharide-mediated signaling pathway | 60 | 18 | 0.000234 | 0.049396 |

**Abbreviations**: N = number of genes in the GO term, DE = number of genes that are differentially methylated in the GO term, P.DE = P value for over representation of the GO term, FDR = false discovery rate from GO analysis

#### Supplementary Table 4: Differentially methylated probes over time following infliximab treatment compared to adalimumab treatment regardless of response

| CpG | chr | pos | Relation to Island | Gene Name | Gene Group | Coefficient | P value |
|---|---|---|---|---|---|---|---|
| cg03446165 | 16 | 3107168 | N_Shore | MMP25 | Body | -0.0004 | 6.78E-10 |
| cg12229367 | 17 | 55822335 | OpenSea | | | -0.0003 | 1.54E-09 |
| cg04790662 | 8 | 82005985 | OpenSea | PAG1 | 5'UTR | -0.0005 | 2.82E-09 |
| cg00508575 | 12 | 90050967 | OpenSea | ATP2B1 | TSS1500 | -0.0004 | 4.91E-09 |
| cg00561739 | 5 | 1807733 | OpenSea | NDUFS6 | Body | -0.0004 | 6.35E-09 |
| cg07273698 | 2 | 46636809 | OpenSea | | | -0.0003 | 1.10E-08 |
| cg19351166 | 2 | 209133632 | S_Shelf | PIKFYVE | 5'UTR | -0.0004 | 1.49E-08 |
| cg19729645 | 17 | 72007145 | OpenSea | | | -0.0003 | 1.73E-08 |
| cg20958086 | 1 | 160759689 | OpenSea | | | -0.0004 | 1.76E-08 |
| cg13508449 | 2 | 109843185 | OpenSea | SH3RF3 | Body | -0.0004 | 3.00E-08 |
| cg23902264 | 1 | 241815413 | OpenSea | WDR64 | TSS200 | -0.0005 | 4.21E-08 |
| cg10230190 | 1 | 85405081 | OpenSea | MCOLN2 | Body | 0.0004 | 5.49E-08 |
| cg02451866 | 10 | 71251604 | OpenSea | TSPAN15 | Body | -0.0003 | 8.29E-08 |

### Supplementary Table 5: Differentially methylated probes with shared associations to other common traits in the EWAS catalog

| CpG | Gene | Trait |
|---|---|---|
| cg16395997 | WDR8 | C-reactive protein, Alcohol consumption per day, Body mass index |
| cg16736826 | EDN2 | Tobacco smoking, Serum low-density lipoprotein cholesterol, Smoking, Sex |
| cg19270739 | | Cleft palate vs cleft lip, Age |
| cg22448090 | KIF21B | C-reactive protein |
| cg22787719 | | Primary Sjogrens syndrome |
| cg23172671 | | C-reactive protein, Body mass index, Total serum IgE, HIV infection, Rheumatoid arthritis, CCL8 protein levels (SeqId = 13748-4), INHBC protein levels (SeqId = 15686-49) |
| cg26227957 | KIAA0090 | C-reactive protein, Alcohol consumption per day, Follicle stimulating hormone, Chronic kidney disease, estimated glomerular filtration rate (eGFR), very early preterm birth (<28 weeks) |
| cg02068690 | DTNB | Primary Sjogrens syndrome |
| cg02560388 | | Body mass index, Primary Sjogrens syndrome, Smoking, Age, Rheumatoid arthritis |
| cg02749511 | | HIV infection, Inflammatory bowel disease |
| cg13293266 | | Ageing |
| cg14210119 | | Ageing |
| cg15705813 | | Alcohol consumption per day |
| cg19351166 | PIKFYVE | Serum high-density lipoprotein cholesterol |
| cg23371680 | | Rheumatoid arthritis, Android tissue mass, Android total mass |
| cg08996521 | CISH | Age |
| cg09610644 | BDH1 | C-reactive protein, Sex, Chronic kidney disease, alcohol withdrawal recovery, estimated glomerular filtration rate (eGFR), very early preterm birth (<28 weeks) |
| cg10324158 | CLEC3B | Primary Sjogrens syndrome, Sex |
| cg12992827 | | C-reactive protein, Alcohol consumption per day, Body mass index, Inflammatory bowel disease, Growth differentiation factor 15, Rheumatoid arthritis, C-Reactive Protein, All cause mortality |
| cg17572056 | OSTalpha | C-reactive protein, HIV infection |
| cg18513344 | MUC4 | C-reactive protein, Body mass index |
| cg22505962 | CLEC3B | Cleft palate vs cleft lip |
| cg26283496 | | Ageing |
| cg06151145 | SLAIN2 | High-density lipoprotein cholesterol, HIV infection, Ageing |
| cg01538969 | DHX16 | C-reactive protein, Alcohol consumption per day |
| cg03149958 | | Alcohol consumption per day, Age, Smoking, HIV infection |
| cg03546163 | FKBP5 | Alcohol consumption per day, Chronic kidney disease, Body mass index, Waist circumference, Body mass index change, All cause mortality, CBLN4 protein levels (SeqId = 5688-65) |
| cg03940776 | SYNJ2 | Body mass index, Primary Sjogrens syndrome, C-Reactive Protein, Age |
| cg15954235 | SPDEF | Sex |

| | | |
|---|---|---|
| cg17501210 | RPS6KA2 | C-reactive protein, Body mass index, Serum high-density lipoprotein cholesterol, Serum triglycerides, Inflammatory bowel disease, Sex, Ulcerative colitis, schizophrenia, All cause mortality, hepatic fat, PZP protein levels (SeqId = 6580-29) |
| cg25114611 | FKBP5;LOC285847 | Tobacco smoking, Lupus nephritis vs lupus without nephritis, Inflammatory bowel disease, Primary Sjogrens syndrome, Smoking, Sex, Rheumatoid arthritis, All cause mortality, Acute myocardial infarction |
| cg25325512 | PIM1 | C-reactive protein, Primary Sjogrens syndrome, Sex, All cause mortality |
| cg02729030 | IMMP2L | Primary Sjogrens syndrome, Age |
| cg26804423 | ICA1 | C-reactive protein, Body mass index, Serum high-density lipoprotein cholesterol, Inflammatory bowel disease, HIV infection, Glycoprotein acetyls, schizophrenia, alcohol withdrawal recovery, CD300A protein levels (SeqId = 5630-48) |
| cg05279866 | | C-reactive protein, Sex |
| cg13821176 | TRIB1 | Serum high-density lipoprotein cholesterol |
| cg16292768 | CLU | Primary Sjogrens syndrome, early preterm birth (<28 weeks) |
| cg02716826 | SUGT1P1;AQP3 | C-reactive protein, Body mass index, Inflammatory bowel disease, Chronic kidney disease, Waist Circumference |
| cg13661827 | | Primary Sjogrens syndrome |
| cg14012059 | LRSAM1 | HIV infection, Rheumatoid arthritis |
| cg14989316 | LOC283050 | Primary Sjogrens syndrome, Rheumatoid arthritis |
| cg19137806 | INPP5A | Alcohol consumption per day |
| cg19588519 | | C-reactive protein, HIV infection, Sex, Inflammatory bowel disease |
| cg23761815 | SLC29A3 | C-reactive protein, Sex, schizophrenia |
| cg25580581 | | HIV infection, Primary Sjogrens syndrome, Ageing, Body mass index |
| cg01218206 | SIK3 | Smoking |
| cg10437839 | PPP2R5B | Schizophrenia |
| cg12535090 | V2 | C-reactive protein, TEX29 protein levels (SeqId = 10557-6) |
| cg18518074 | EHD1 | Schizophrenia |
| cg00159243 | SELPLG | C-reactive protein, Schizophrenia, Primary Sjogrens syndrome, CBLN4 protein levels (SeqId = 5688-65) |
| cg01871427 | | Primary Sjogrens syndrome |
| cg04255937 | SETD1B | C-reactive protein, Smoking, Rheumatoid arthritis, schizophrenia |
| cg07986378 | ETV6 | Tobacco smoking, Smoking |
| cg12053291 | SCARB1 | C-reactive protein, HIV infection |
| cg13150925 | | Sex |
| cg14849578 | SCARB1 | C-reactive protein, HIV infection |
| cg02003183 | CDC42BPB | C-reactive protein, Alcohol consumption per day, Educatio l attainment, Primary Sjogrens syndrome, Rheumatoid arthritis, Ageing, Inflammatory bowel disease, CBLN4 protein levels (SeqId = 5688-65) |
| cg13976502 | C14orf43 | Alcohol consumption per day, Tobacco smoking, Smoking, Diet Quality Alter tive Healthy Eating Index 2010 (AHEI-2010) |
| cg19769147 | PACS2 | C-reactive protein, Alcohol consumption per day, Schizophrenia, Rheumatoid arthritis, Inflammatory bowel disease |
| cg22908922 | PACS2 | C-reactive protein, Alcohol consumption per day, Chronic kidney disease, EBAG9 protein levels (SeqId = 19323-1) |
| cg25132241 | FBLN5 | C-reactive protein, Inflammatory bowel disease |
| cg09018739 | CPNE2 | C-reactive protein, Alcohol consumption per day, Body mass index |
| cg09481056 | ABCA3 | C-reactive protein, Alcohol consumption per day, Rheumatoid arthritis |
| cg23669118 | C16orf38 | HIV infection |

| | | |
|---|---|---|
| cg02976843 | RAP1GAP2 | C-reactive protein, Alcohol consumption per day, HIV infection |
| cg09929238 | RPTOR | Alcohol consumption per day |
| cg15090877 | P4HB | Sex, LDL cholesterol |
| cg17781958 | BAHCC1 | HIV infection |
| cg24129923 | CHD3 | Alcohol consumption per day |
| cg25616160 | | Body mass index |
| cg00711496 | C19orf76;PRMT1 | Smoking, Free cholesterol to total lipids ratio in large HDL, Glycoprotein acetyls |
| cg01799015 | PALM | Alcohol consumption per day, Primary Sjogrens syndrome, Inflammatory bowel disease, Chronic kidney disease |
| cg10604476 | ICAM5 | Primary Sjogrens syndrome, Sex, Ageing, All cause mortality |
| cg17417856 | PRMT1;C19orf76 | Alcohol consumption per day, Tobacco smoking, Smoking, Educatio l attainment, Age |
| cg22910295 | ICAM5 | Primary Sjogrens syndrome, Sex |
| cg26470501 | BCL3 | C-reactive protein, Alcohol consumption per day, Body mass index, Smoking, Inflammatory bowel disease, Educatio l attainment, Primary Sjogrens syndrome, Rheumatoid arthritis, lung cancer, All cause mortality |
| cg08507178 | PIM3 | C-reactive protein, Arsenic exposure |
| cg09349128 | | Body mass index, Inflammatory bowel disease, Primary Sjogrens syndrome, Growth differentiation factor 15, PACAP protein levels (SeqId = 16322-10), orofacial cleft (cleft lip or cleft lip and palate) |
| cg17237086 | MKL1 | C-reactive protein, Alcohol consumption per day, HIV infection |
| cg24162801 | C1orf21 | Haemoglobin, EGLN1 protein levels (SeqId = 9901-28) |
| cg06291107 | BLCAP | Arsenic exposure, Inflammatory bowel disease |
| cg13572071 | TCEA3 | Age |
| cg12670943 | POLDIP3 | Rheumatoid arthritis |
| cg00053916 | | Inflammatory bowel disease |
| cg07058694 | ATXN2 | Inflammatory bowel disease |
| cg08943045 | | Inflammatory bowel disease, FCRL1 protein levels (SeqId = 5728-60) |
| cg17325206 | | alcohol withdrawal recovery |
| cg27216853 | CYS1 | alcohol withdrawal recovery, CHRDL2 protein levels (SeqId = 6086-15) |
| cg23608915 | MAP4K4 | alcohol withdrawal recovery |
| cg09842038 | CYS1 | alcohol withdrawal recovery |
| cg07534027 | SAG | alcohol withdrawal recovery |
| cg09310636 | NRDE2 | alcohol withdrawal recovery |
| cg09197509 | | alcohol withdrawal recovery |
| cg01211124 | NUPL1 | alcohol withdrawal recovery |
| cg12716582 | | alcohol withdrawal recovery |
| cg05673431 | KSR1 | alcohol withdrawal recovery, PACAP protein levels (SeqId = 16322-10) |
| cg21800396 | ACSM6 | malathion |
| cg27064284 | MIR1273H | metolachlor |
| cg18871648 | ELMSAN1 | Waist Circumference, Body mass index |
| cg19748455 | LOC100996291 | Body mass index, PACAP protein levels (SeqId = 16322-10) |
| cg22698218 | CNOT4 | very early preterm birth (<28 weeks) |
| cg00519063 | GCLM | very early preterm birth (<28 weeks), early preterm birth (<28 weeks) |
| cg24298539 | | very early preterm birth (<28 weeks) |
| cg21737354 | TSPAN15 | HNF4A protein levels (SeqId = 10041-3) |
| cg26081786 | | HNF4A protein levels (SeqId = 10041-3) |

| | | |
|---|---|---|
| cg18931885 | | HNF4A protein levels (SeqId = 10041-3), R SE6 protein levels (SeqId = 5646-20), ARHGAP36 protein levels (SeqId = 6289-78), VAV3 protein levels (SeqId = 9830-109) |
| cg20292908 | | NTM protein levels (SeqId = 10907-116), OPCML protein levels (SeqId = 15622-13) |
| cg04609197 | GHRH | COL11A2 protein levels (SeqId = 11278-4) |
| cg21489638 | LINC-PINT | IL5RA protein levels (SeqId = 13686-2) |
| cg02907982 | ADGRD1 | FGF19 protein levels (SeqId = 13724-27) |
| cg11424260 | A PC7 | GLIPR2 protein levels (SeqId = 15522-2) |
| cg00840791 | | PACAP protein levels (SeqId = 16322-10) |
| cg18353028 | CYP7B1 | PACAP protein levels (SeqId = 16322-10) |
| cg15001240 | | PACAP protein levels (SeqId = 16322-10), GALNT2 protein levels (SeqId = 5764-4) |
| cg11902329 | | PACAP protein levels (SeqId = 16322-10) |
| cg03262246 | CDKN2AIPNL | MLN protein levels (SeqId = 5631-83) |
| cg12174507 | RCSD1 | TMX3 protein levels (SeqId = 5654-70), EPO protein levels (SeqId = 5813-58) |
| cg01467435 | GTDC1 | FCRL1 protein levels (SeqId = 5728-60) |
| cg03172765 | PSMD1 | FTH1 protein levels (SeqId = 5934-1) |
| cg22417925 | TOP1MT | FTH1 protein levels (SeqId = 5934-1) |
| cg21452109 | | WSCD2 protein levels (SeqId = 6274-15) |
| cg03698766 | | RPN1 protein levels (SeqId = 6458-6) |
| cg00156232 | VKORC1L1 | ENPP5 protein levels (SeqId = 6556-5) |
| cg10967866 | INPP5A | FAIM3 protein levels (SeqId = 6574-11) |
| cg04940901 | D H1 | VAV3 protein levels (SeqId = 9830-109) |

### Supplementary Table 6: Differentially methylated probes at baseline associated with both drug concentration at week 14 and primary non-response

| CpG | chr | pos | Relation to Island | Gene Name | Gene Group | Drug level Coefficient | Drug level p value | PNR Coefficient | PNR P value |
|---|---|---|---|---|---|---|---|---|---|
| cg27216853 | 2 | 10205672 | OpenSea | CYS1 | Body | -0.0371 | 8.79E-12 | 0.0245 | 2.38E-10 |
| cg23606775 | 1 | 9790616 | Island | CLSTN1 | Body | -0.0220 | 1.44E-11 | 0.0133 | 2.62E-08 |
| cg18138532 | 10 | 12078337 | OpenSea | UPF2 | 5'UTR | -0.0273 | 1.92E-11 | 0.0157 | 8.51E-08 |
| cg09842038 | 2 | 10205716 | OpenSea | CYS1 | Body | -0.0361 | 5.95E-11 | 0.0245 | 1.26E-09 |
| cg16217672 | 10 | 75843201 | OpenSea | VCL | Body | -0.0264 | 1.76E-10 | 0.0167 | 8.61E-08 |
| cg01499619 | 5 | 179205250 | OpenSea | | | -0.0249 | 3.73E-10 | 0.0170 | 2.27E-09 |
| cg10705487 | 5 | 179108352 | S_Shore | CBY3 | TSS1500 | -0.0217 | 3.73E-10 | 0.0139 | 4.72E-08 |
| cg22090525 | 10 | 12078334 | OpenSea | UPF2 | 5'UTR | -0.0254 | 4.48E-10 | 0.0177 | 1.33E-09 |
| cg10100117 | 1 | 179843111 | N_Shelf | TOR1AIP2 | 5'UTR | -0.0269 | 5.82E-10 | 0.0183 | 8.79E-09 |
| cg16395997 | 1 | 3562798 | S_Shore | WDR8 | Body | -0.0369 | 7.87E-10 | 0.0241 | 2.02E-08 |
| cg23608915 | 2 | 102333795 | OpenSea | MAP4K4 | Body | -0.0312 | 9.42E-10 | 0.0215 | 6.26E-09 |
| cg09439276 | 16 | 66554515 | OpenSea | TK2 | Body | -0.0222 | 2.41E-09 | 0.0156 | 6.38E-09 |
| cg23669118 | 16 | 1538347 | S_Shore | C16orf38 | 1stExon | -0.0379 | 2.63E-09 | 0.0245 | 4.54E-08 |
| cg03070999 | 12 | 6745630 | OpenSea | LPAR5 | TSS1500 | -0.0218 | 5.56E-09 | 0.0151 | 4.91E-08 |
| cg04326337 | 20 | 36664969 | S_Shelf | RPRD1B | Body | -0.0354 | 5.69E-09 | 0.0250 | 4.44E-08 |
| cg03546163 | 6 | 35654363 | N_Shore | FKBP5 | 5'UTR | 0.0669 | 1.57E-08 | -0.0488 | 1.50E-08 |
| cg02897385 | 5 | 142528481 | OpenSea | ARHGAP26 | Body | 0.0337 | 2.57E-08 | -0.0258 | 4.46E-08 |
| cg18931885 | 10 | 112100739 | OpenSea | | | -0.0257 | 3.09E-08 | 0.0190 | 1.11E-08 |
| cg03262246 | 5 | 133749006 | S_Shore | CDKN2AIPNL | TSS1500 | -0.0189 | 5.23E-08 | 0.0135 | 3.86E-08 |
| cg13661827 | 9 | 111885602 | S_Shelf | | | 0.0240 | 8.53E-08 | -0.0173 | 8.60E-08 |

#### Supplementary Table 7: Differentially methylated probes associated with anti-TNF primary non-response over time

| CpG | chr | pos | Relation to Island | Gene name | Gene group | Coefficient | P value |
|---|---|---|---|---|---|---|---|
| cg07839457 | 16 | 57023022 | N_Shore | NLRC5 | TSS1500 | -0.0007 | 1.93E-13 |
| cg11047325 | 17 | 76354934 | Island | SOCS3 | Body | -0.0007 | 3.70E-11 |
| cg15022400 | 15 | 45028161 | OpenSea | TRIM69 | TSS1500 | -0.0003 | 3.33E-09 |
| cg25867318 | 17 | 40494745 | OpenSea | STAT3 | Body | -0.0004 | 6.06E-08 |
| cg08950751 | 11 | 67255492 | OpenSea | AIP | Body | -0.0003 | 6.82E-08 |

#### Supplementary Figure 1: Sample flow through the study

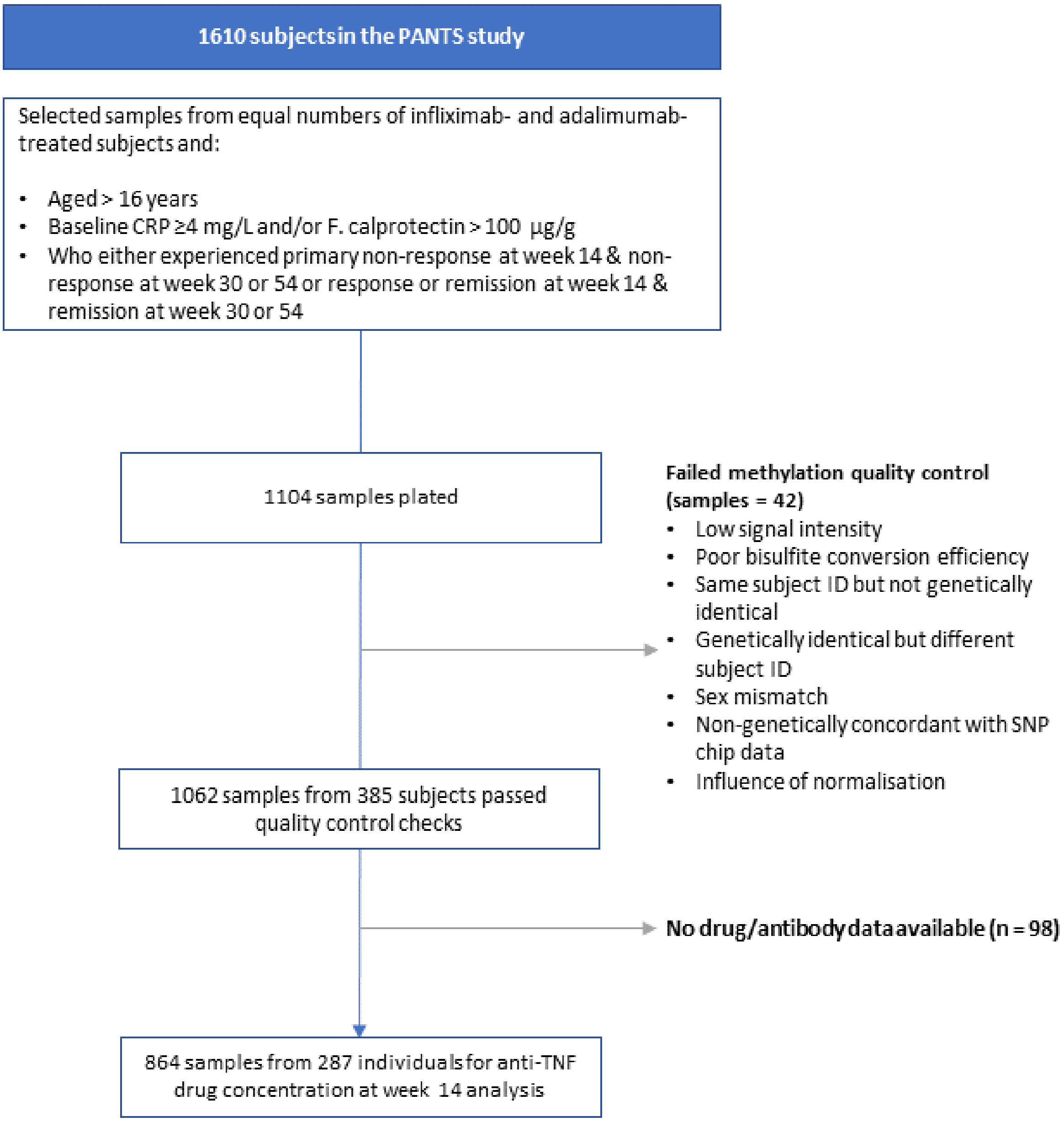

#### Supplementary Figure 2: Density plot of anti-TNF drug concentration at week 14 stratified by primary non-response

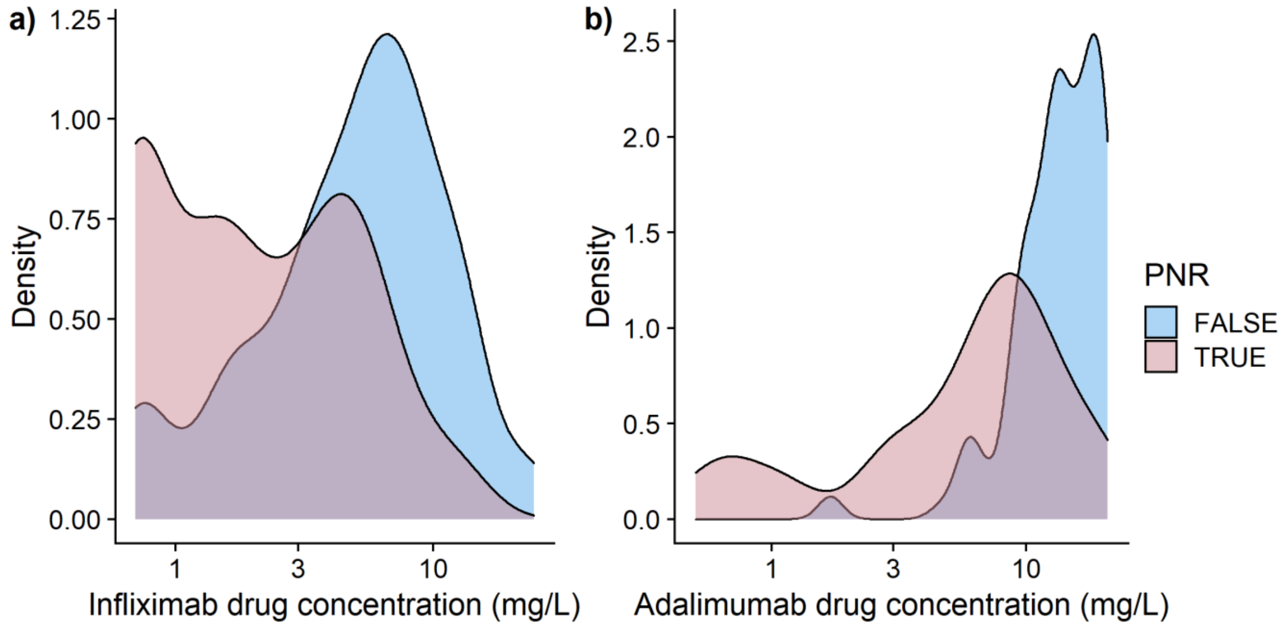

a) Infliximab drug concentration at week 14 stratified by primary non-response status b) adalimumab drug concentration at week 14 stratified by primary non-response status. Abbreviations: PNR = primary non-response

#### Supplementary Figure 3: Change in DNA methylation smoking score over 54 weeks

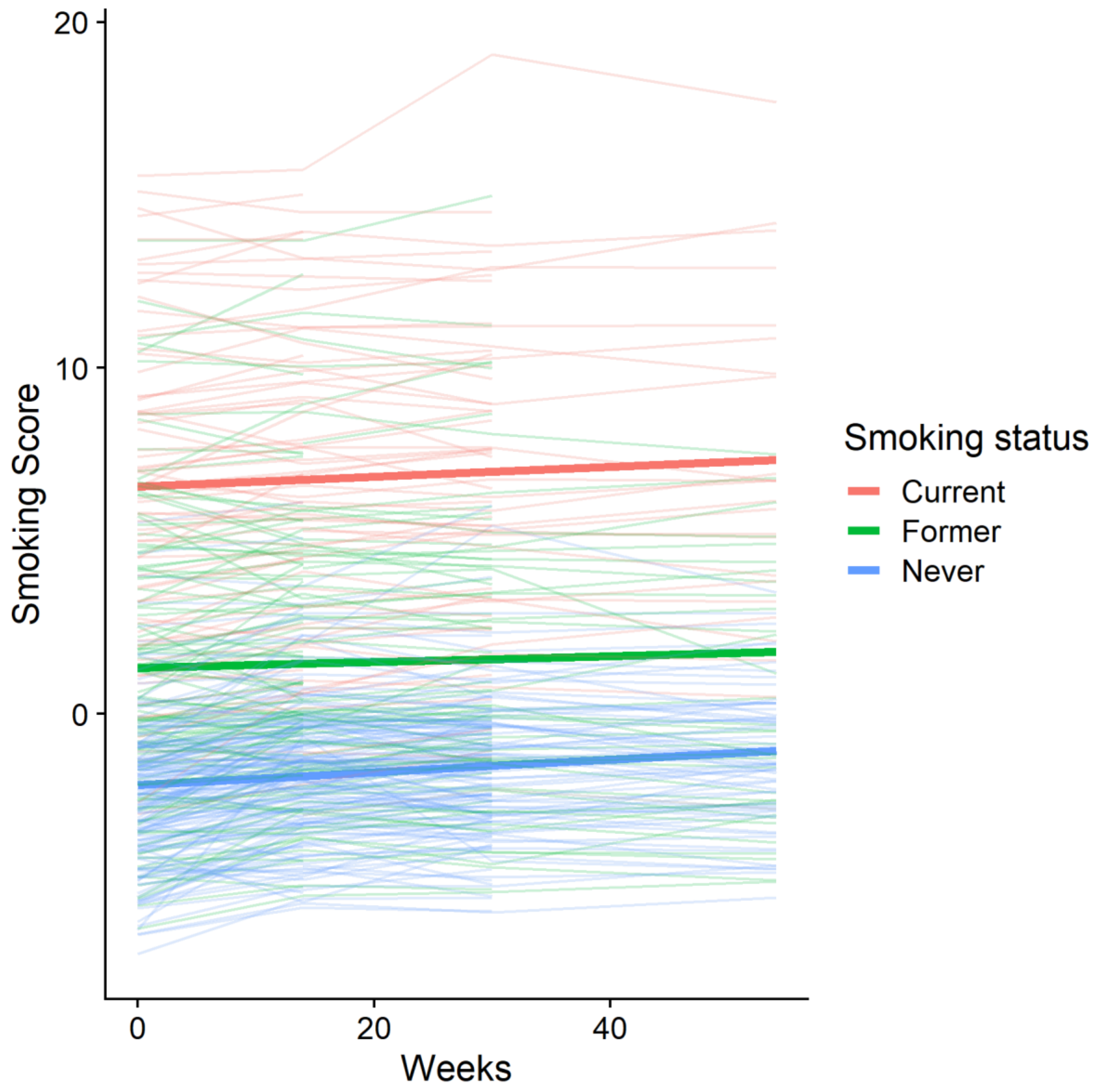

Predicted smoking scores in solid lines and un-corrected smoking scores in faded lines. Red represents smoking score of current smokers, green of former smokers and blue of never smokers.

#### Supplementary Figure 4: Change in epigenetic age over 54 weeks

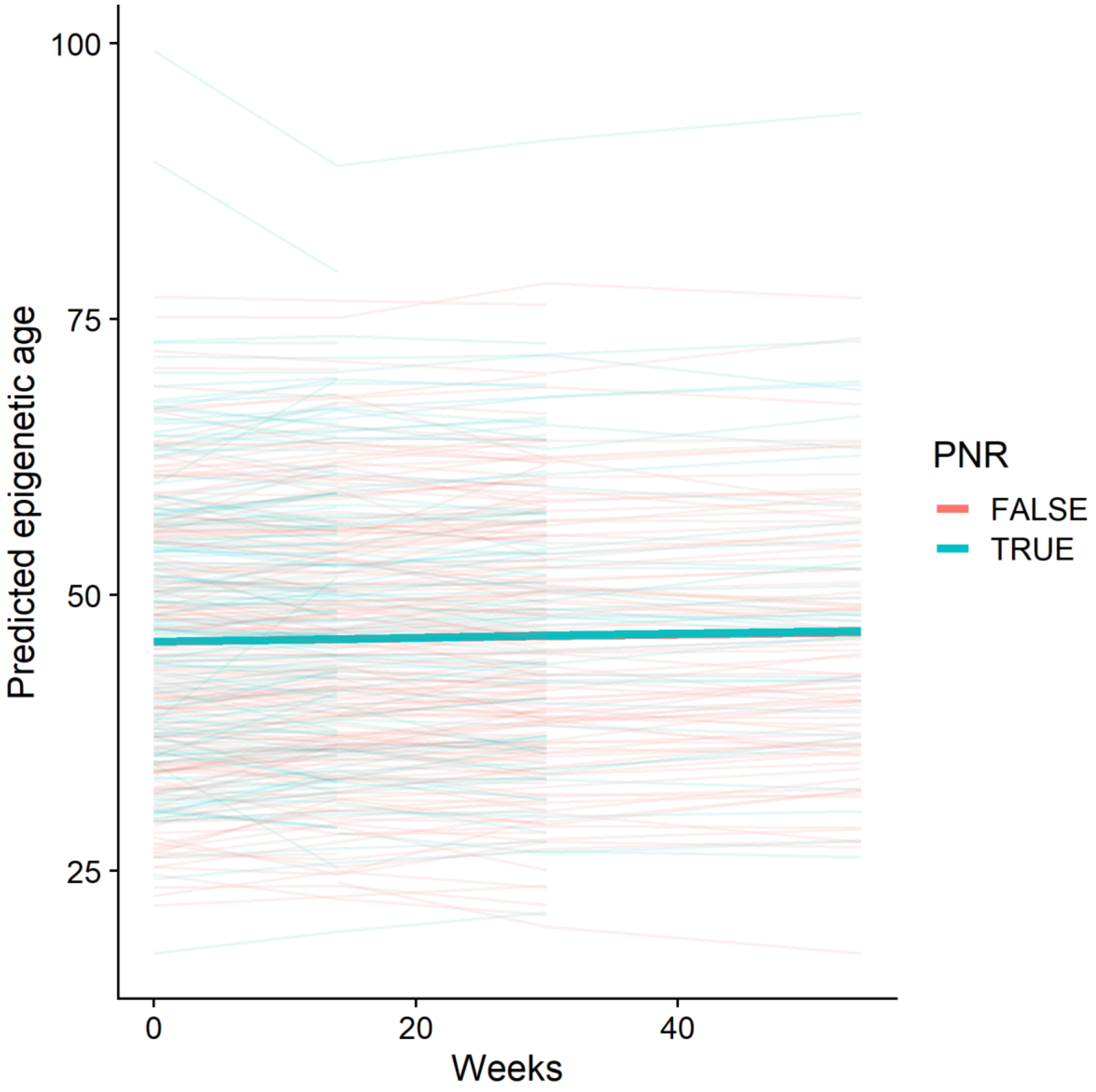

Predicted epigenetic age in solid lines and un-adjusted epigenetic age in faded lines. Turquoise represents those who experienced primary non-response and orange those who did not. PNR = primary non-response.

#### Supplementary Figure 5: Genomic location of differentially methylated probes associated with anti-TNF treatment over time

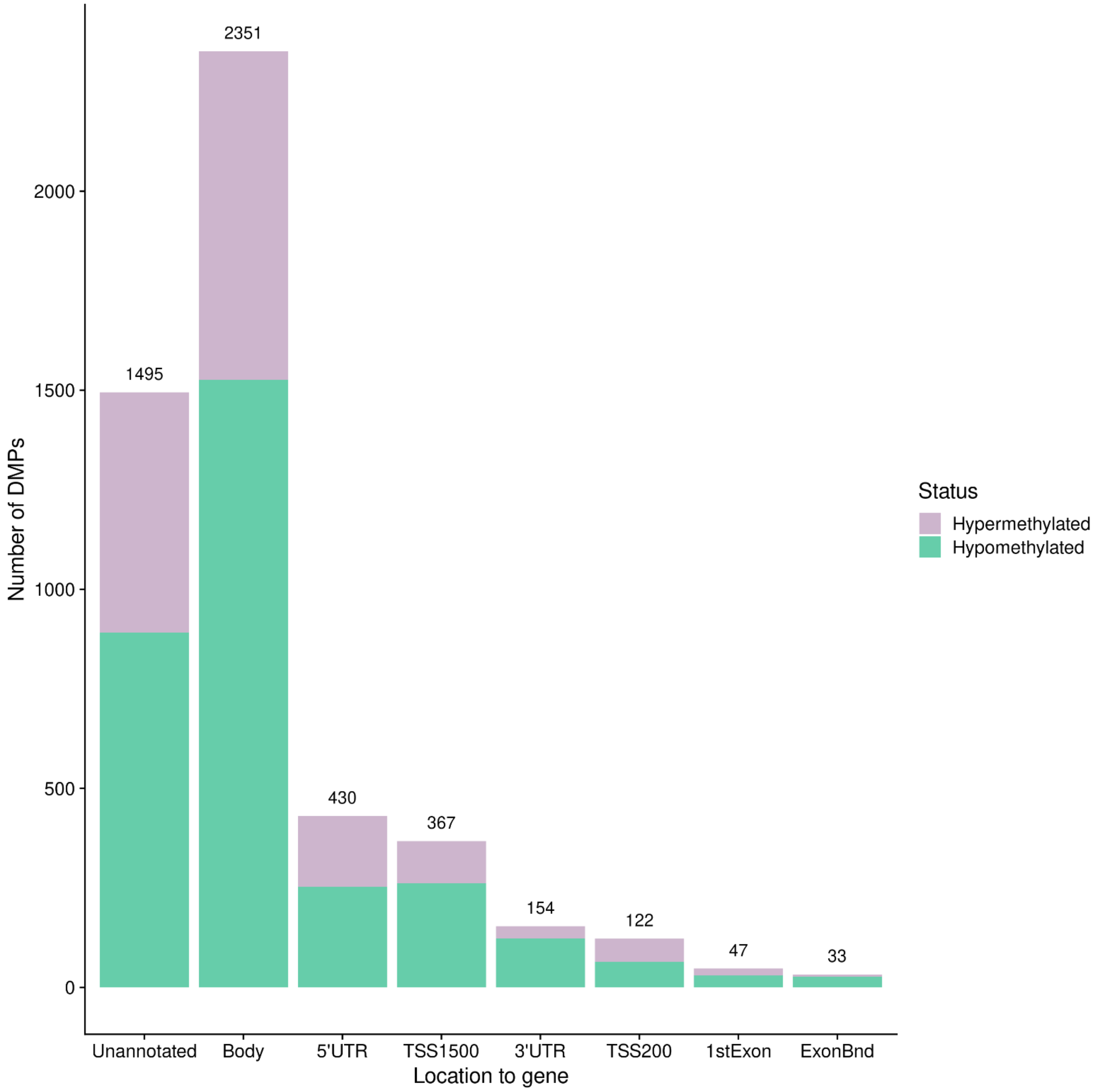

Genomic locations as provided by Illumina. Green represents hypomethylated DMPs, purple represents hypermethylated DMPs. Abbreviations: DMPs = differentially methylated positions, UTR = untranslated region, TSS = transcription start site, ExonBnd = exon boundary.

#### Supplementary Figure 6: Bar chart of the top 20 Gene Ontology terms of differentially methylated probes associated with anti-TNF treatment

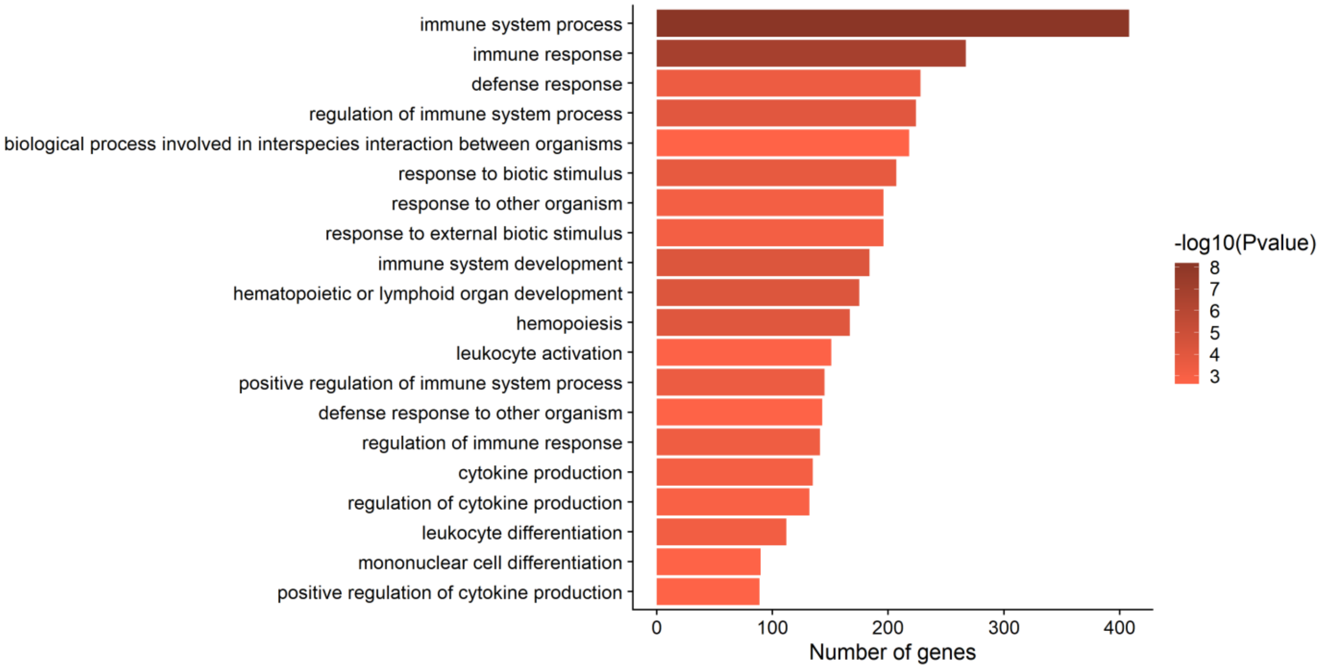
